# The QuantuMDx Q-POC™ SARS-CoV-2 RT-PCR assay for rapid detection of COVID-19 at point-of-care: preliminary evaluation of a novel technology

**DOI:** 10.1101/2021.07.12.21260119

**Authors:** Jessica Caffry, Matthew Selby, Katie Barr, George Morgan, David McGurk, Philip Scully, Catherine Park, Anna-Maria Caridis, Emily Southworth, Jack Morrison, David J Clark, Nickolas Eckersely, Elisabetta Groppelli, Daniela E. Kirwan, Irene Monahan, Yolanda Augustin, Colin Toombs, Tim Planche, Henry M Staines, Sanjeev Krishna

**Author notes:** Corresponding authors: Sanjeev Krishna, Henry Staines. Equal contribution: Jessica Caffry & Matthew Selby.

## Abstract

**Background:** Accurate, affordable, and rapid point-of-care (PoC) diagnostics are critical to the global control and management of the COVID-19 pandemic. The current standard for accurate diagnosis of SARS-CoV-2 is laboratory-based reverse transcription polymerase chain reaction (RT-PCR). Here, we report a preliminary prospective performance evaluation of the QuantuMDx Q-POC™ SARS CoV-2 RT-PCR assay.

**Methods:** Between November 2020 and March 2021, we obtained 49 longitudinal nose and throat swabs from 29 individuals hospitalised with RT-PCR confirmed COVID-19 at St George’s NHS Foundation Trust, London (UK). In addition, we obtained 101 mid nasal swabs from healthy volunteers in June 2021. We then used these samples to evaluate the Q-POC™ SARS-CoV-2 RT-PCR assay. The primary analysis was to compare the sensitivity and specificity of the Q-POC™ test against a reference laboratory-based RT-PCR assay.

**Results:** The overall sensitivity of the Q-POC™ test compared with the reference test was 96.88% (83.78%-99.92% CI) for a cycle threshold (Ct) cut-off value for the reference test of 35 and 80.00% (64.35% to 90.95% CI) without altering the reference test’s Ct cut-off value of 40.

**Conclusions:** The Q-POC™ test is a sensitive, specific and rapid point-of-care test for SARS-CoV-2 at a reference Ct cut-off value of 35. The Q-POC™ test provides an accurate and affordable option for RT-PCR at point-of-care without the need for sample pre-processing and laboratory handling. The Q-POC™ test would enable rapid diagnosis and clinical triage in acute care and other settings.

## Introduction

Severe acute respiratory syndrome coronavirus 2 (SARS-CoV-2) is an enveloped, positive sense, single-stranded RNA viruses of the genus betacoronavirus that caused the corona virus disease 2019 (COVID-19) pandemic with devastating public health impact. As of 24^th^ June 2021, there have been more than 180 million individuals infected and almost 4 million deaths globally (1). The first diagnostic assays for this disease were developed shortly after the initial publication of the viral genome in January 2020 (2). These were nucleic acid amplification tests (NAATs) based on reverse transcriptase polymerase chain reaction (RT-PCR), where target viral RNA sequences are converted to DNA and exponentially amplified to allow detection with fluorescent probes. The lower the concentration of target RNA sequence in a clinical sample, the higher the cycle threshold (Ct) value at which fluorescence is detectable.

Most convenient assays use upper respiratory (*e*.*g*. nasopharyngeal or oropharyngeal or nose and throat) swab samples to detect SARS-CoV-2 RNA and are highly sensitive (e.g. (3)). Viral load is highest in the upper respiratory tract in the first week of illness (4). However, standard laboratory-based RT-PCR assays are relatively complex and time consuming, requiring highly trained personnel and expensive equipment based in a centralised laboratory. They take several hours to perform in batches, and, when time is added for sample transport, results often take 24 hours or more to be reported. This can result in delayed diagnosis, which can complicate clinical triage.

Point-of-care (PoC) diagnostics were identified as one of eight key research priorities to tackle the pandemic (5). Rapid, PoC or near-patient testing for active infections confers several benefits over laboratory-based testing including faster triage, which in turn aids infection control and clinical management. Rapid PoC tests can also enable effective community testing and alleviate pressure on overburdened centralised labs. While lateral flow-based rapid antigen tests (RDTs) are now in widespread use, they are less sensitive than NAATs (6). Thus, development of rapid PoC NAAT-based tests are a priority, especially for use when immediate actions are needed for positive cases such as in accident and emergency departments, pre-surgical or chemotherapy day unit admissions, care homes, airports, prisons and low resource healthcare settings. Several versions of PoC NAATs have been developed (7-10), but all have one or more limitations including long sample-to-result times, complexity (steps required) and cost.

To address the urgent need for a rapid and sensitive COVID-19 diagnostic, QuantuMDx have repurposed their RT-PCR PoC diagnostics platform, the Q-POC™, to run a SARS-CoV-2 assay. Here, we present a detailed description of this technology and a preliminary prospective performance evaluation of Q-POC™ compared with validated laboratory-based RT-PCR assays.

## Methods

### Ethical statement

Ethical approval for clinical sample use at SGUL was provided by the Institutional Review Board as part of the “Development and Assessment of Rapid Testing for SARS-CoV-2 outbreak” (DARTS) study, sponsored by St George’s Hospital NHS Foundation Trust (Integrated Research Application System project ID: 282104; South Central - Oxford C Research Ethics Committee reference: 20/SC/0171; registered at clinicaltrials.gov NCT04351646). All participants for this study were recruited following informed consent.

### Swab samples

Patients with COVID-19 hospitalised at St George’s Hospital were recruited into the prospective study between 19^th^ November 2020 and 3^rd^ March 2021 (inclusive). Prior to recruitment, they were confirmed as positive for SARS-CoV-2 using RT-PCR from nose and throat swabs (in Sigma Virocult®, Corsham, UK) and Roche RNA extraction kits (Magnapure, West Sussex, UK) followed by altona Diagnostics RealStar® SARS-CoV-2 RT-PCR (S and E target genes, Hamburg, Germany) or Roche cobas® SARS-CoV-2 Test (E and ORF target genes). All patients consented to nose and throat swabs being taken. Following recruitment to DARTS, longitudinal nose and throat swab samples were collected, using a Copan swab (Cat number: 503CS01). Samples were stored within 2 hours, without viral transport medium, at -80 C until use. In addition, anonymised mid-nasal swab samples were collected between 15^th^ to the 18^th^ June 2021 (inclusive) from healthy staff volunteers at QuantuMDx following informed consent and tested immediately.

### Q-POC™ testing

The Q-POC™ device is a PoC diagnostics platform, which runs a rapid molecular test in a single use microfluidic cassette (Q-CAS) by incorporating sample processing, DNA amplification (with RT step, if required), and downstream semi-quantitative detection of pathogens in small volumes of sample.

For detection of SARS-CoV-2 by RT-PCR at PoC a flexible mini tip Copan FLOQSwab (designed for nasopharyngeal sampling) was used to take a nose and throat or a mid nasal swab. The tip (fresh or defrosted) was then broken off into a sample tube containing 3 ml of MSwab™ sample collection, transport and preservation medium (Copan Diagnostics, Italy). Following sample tube inversion (x5), a 400 µl aliquot is pipetted into a test cassette, Q-CAS (Figure 1) and sealed in with a cap.

**Figure 1.**
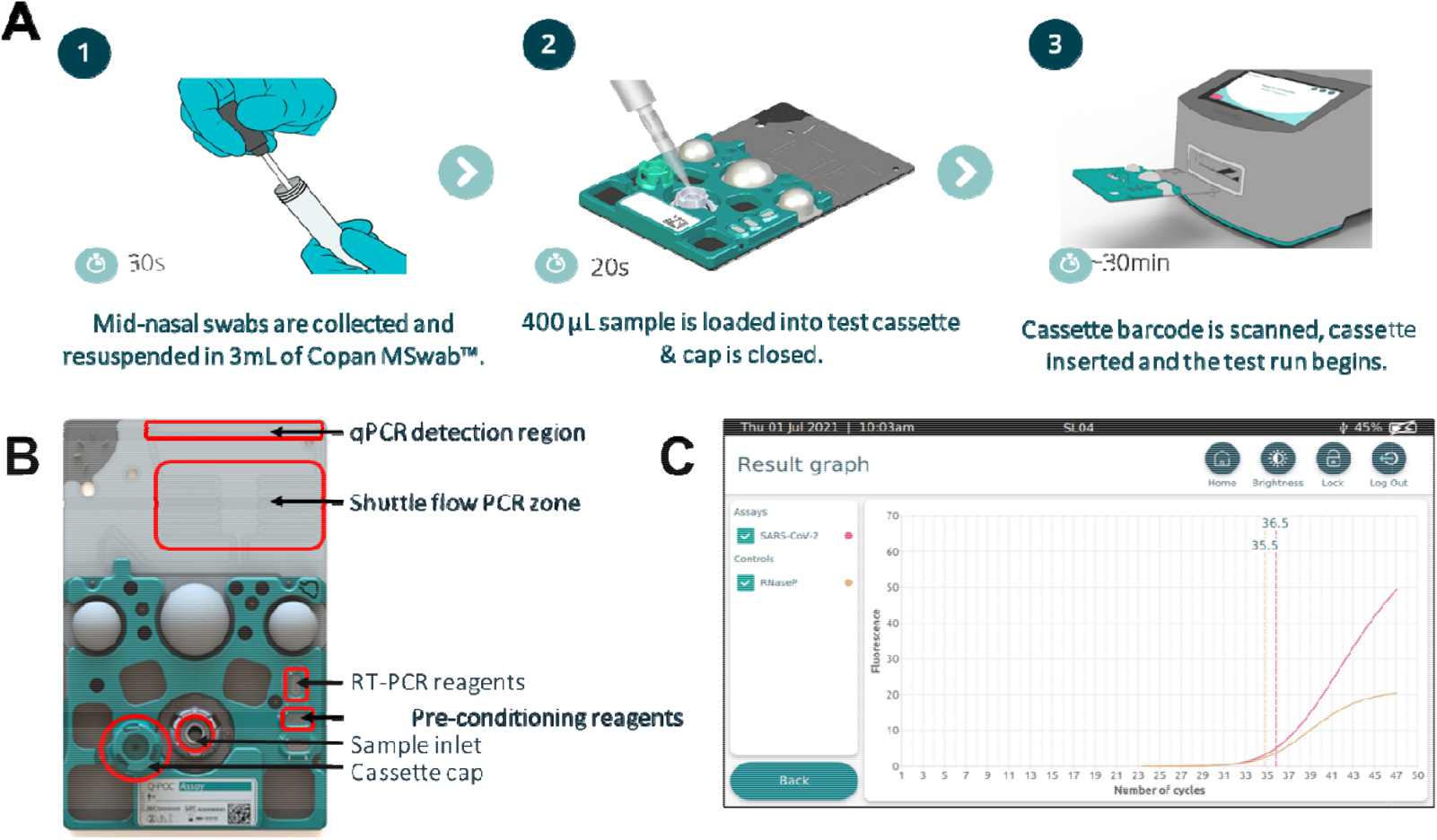
QuantuMDx Q-POC^™^ and Q-CAS, point-of-care diagnostic for detection of SARS-CoV-2 from nasal swab samples. A. Graphic showing the Q-POC^™^/Q-CAS workflow. A midnasal swab is collected and resuspended in Copan MSwab (3 ml). The tube is inverted 5 times before 400 µl is dispensed into the inlet of the cassette using a pipette. The barcode is scanned, Q-POC^™^ identifies the cassette, the cassette is inserted and Q-POC^™^ begins the SARS-CoV-2 assay. A result is displayed on screen in ∼30 minutes. B. Key areas of the Q-CAS cassette. Q-POC^™^ interacts with the self-contained cassette to move the sample from the inlet through the lyophilised reagents and into the shuttle flow PCR region. RT-PCR amplification is detected during shuttling through the detection region. C. Results graph for a positive SARS-CoV-2 sample. The assay results in semi-quantitative PCR curves for SARS-CoV-2 (in red) and sample control, RNaseP (in yellow). This screen is optionally displayed after the main results screen whereby a definitive result is displayed (Positive/Negative/Invalid). Results can be reviewed, printed and uploaded.

The loaded cassette is then scanned and loaded into the Q-POC™ device. Within the Q-CAS, reconstitution of lyophilised RT-PCR reagents, held within sealed ‘Reagent Fuses’ for increased shelf-life, occurs using 50 µl of the swab eluate containing released RNA. Following an RT step, rapid PCR is achieved by shuttling fluid between heating zones (shuttle flow PCR, sfPCR – Fig. 1B). This allows target amplification in approximately 32 minutes. SARS-CoV-2 gene targets (Orf1ab, N and S) and an RNaseP (control) are detected by fluorescence readings taken after each cycle (qPCR reader), for semi-quantitative analysis. The use of individual heating zones combined with shuttling of the fluid, unlike the temperature ramping used by conventional PCR, is far more energy efficient allowing for multiple assay runs even when the Q-POC™ is battery operated. The Q-POC™ is front-loading and intuitive, with a simple touch screen interface designed for PoC testing in resource limited settings. It contains all the necessary mechanics, electronics and optics to drive the cassette-based assays, in addition to reading, interpreting and presenting test results. It is portable and operable either on mains power or by an exchangeable battery, which can last up to 4 h. Data are recorded and initially stored on the Q-POC™ within a secured database, with a storage capacity of >1000 patient results. In addition, the Q-POC™ will connect to a specified receipt-style printer. The Q-POC™ operating system runs on an ARM-based processing board integrating connectivity options: GPS, Bluetooth, Wi-Fi and GPRS (2G/3G) connectivity. Operation data are also retained within the Q-POC™ to allow for tracking and trending of instrument performance.

### Laboratory based reference testing

An aliquot (140 µl) of each swab sample in 3 ml of MSwab was used with a QIAamp Viral RNA Mini Kit (Qiagen, Germany) for RNA extraction, as per the manufacturer’s instructions. The CE-marked SARS-CoV-2 RT-PCR Detection Assay (QuantuMDx, UK) was used as the reference test. It is based on TaqMan chemistry and utilises FAM for the detection of the three SARS-CoV-2 loci, Orf1ab, N gene and the S gene and HEX for the detection of the RNase P gene (a specimen and process control).

### Statistical analysis

Operators were not blinded to the status of patients/volunteers from which the samples were taken prior to Q-POC™ testing but did not have access to the results of the subsequent reference testing; however, the Q-POC™ requires no subjective interpretation of a result.

Data were analysed with GraphPad Prism (version 6.07 for Windows). Diagnostic sensitivity, specificity, positive predictive value and negative predictive value were determined, using the laboratory-based RT-PCR as a reference standard (Supplementary data). Samples that were invalid on the PoC platform were not included in the primary sensitivity analysis. The manuscript was prepared in accordance with the EQUATOR Network’s STARD guidelines (Supplementary checklist).

## Results

Swab samples (n = 150) were collected and tested, using the Q-POC™ (Fig. 2). Two cohorts were recruited. In the first, hospitalised patients at St George’s Hospital with an RT-PCR confirmed COVID-19 diagnosis were clinically assessed between the 19^th^ November 2020 and 3^rd^ March 2021 as part of DARTS. The recruits provided longitudinal samples predicted to have a wide range of viral loads thereby provide a challenging sample set for initial evaluation of the Q-POC™. Patients (n = 29) who provided a nose and throat sample, using a mini tip Copan swab, were eligible for inclusion in the Q-POC™ evaluation. One patient’s sample produced an invalid test result and was not retested. This patient provided only one sample so, while the invalid result is included in the analysis (Fig. 2), the patient is not (Table 1). Therefore, complete clinical data paired with laboratory tests were available for 28 patients and 47 samples. Participants formed a convenience series. No adverse events from performing the Q-POC™ or the reference test were recorded.

**Figure 2.**
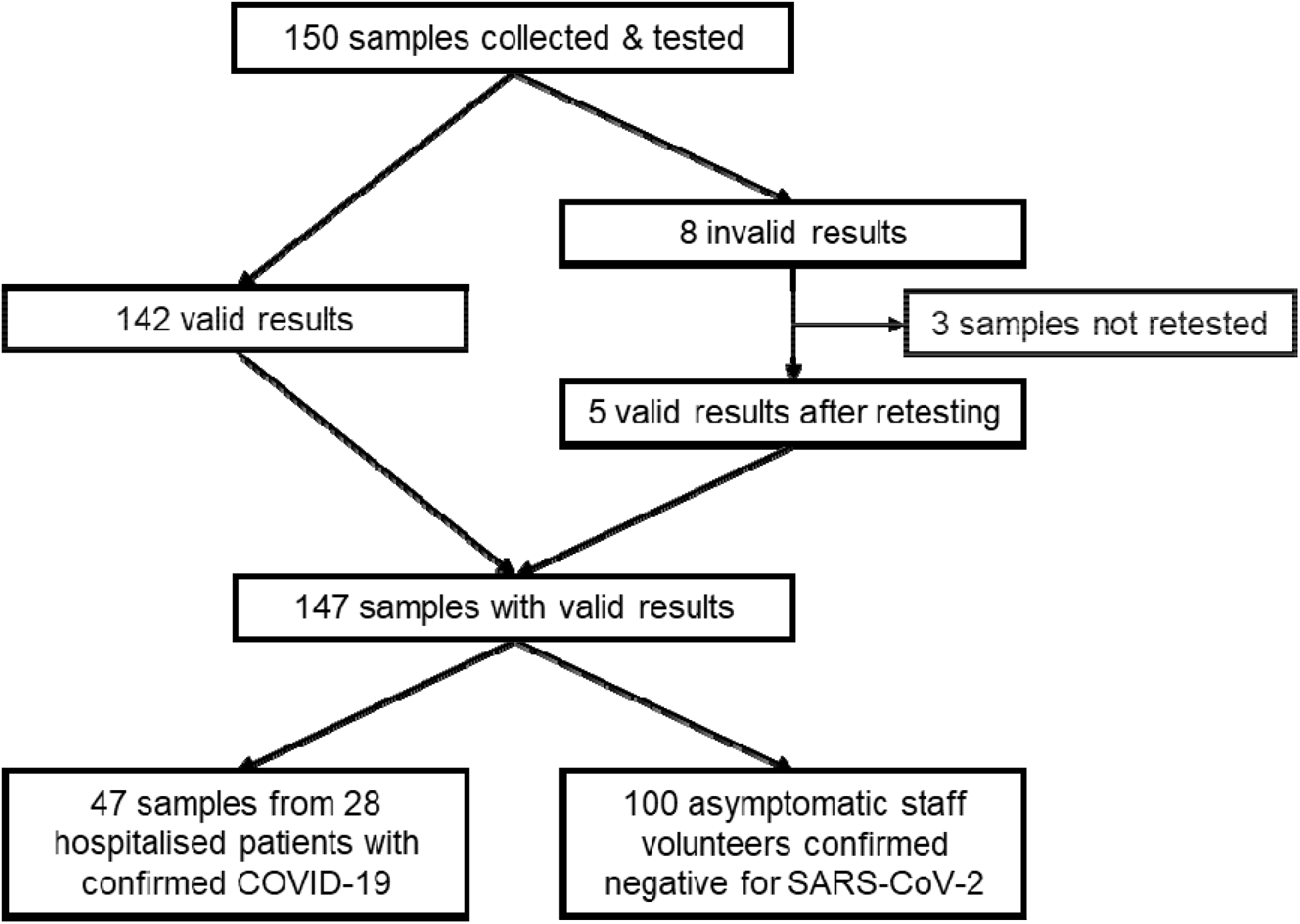
Study design.

**Table 1.** Patient demographics.

| Demographic | N (%) |
| --- | --- |
| Total number of patients included in study | 28 |
| Male Sex | 19 (68%) |
| Median Age (Years) | 68 |
| Symptomatic at time of study enrolment | 15 (54%) |
| WHO Ordinal Symptom Severity Scale (OSSS) Baseline Enrolment* | WHO OSSS 4 = 23 (82%)<br>WHO OSSS 5 = 4 (14%)<br>WHO OSSS 6 = 1 (4%) |

\* WHO OSSS 4 = Hospitalised, mild disease, no oxygen therapy; WHO OSSS 5 = Hospitalised, oxygen mask or nasal prongs; WHO OSSS 6 = Hospitalised, non-invasive ventilation/high flow oxygen

The median age of COVID-19 positive study participants included in the analysis was 64.5 (IQR 51-75), 19 (68%) were male, 15 (54%) were symptomatic and the majority (23; 82%) had mild disease at enrolment (Table 1). In the second cohort, 101 mid nasal Copan swab samples were collected from healthy volunteer staff to provide a SARS-CoV-2 negative dataset. Of these, 1 sample returned an invalid test result and was not retested. Therefore, test data for further analysis were available for 100 samples from this cohort (Fig. 2). Of the total 155 sample runs on the Q-POC™ there were 8 invalid runs, 5 of which were retested to produce valid results, and 3 of which were not retested as the sample was destroyed (Fig. 2). This gives an invalid rate over the study period of 5.2%.

Figure 3 presents the Ct values from the reference test used with longitudinal nose and throat swab samples from COVID-19 positive participants. As predicted, this sample set provided a wide range of viral loads (using Ct values as a basic proxy), including multiple samples between Ct values of 30 and 40. Included in the graph are the available Ct data from the hospital test undertaken prior to enrolment (for 22 of the 28 participants). The day the swab was taken for this test was used as day 0. Two different validated assays (see Methods) were used at St George’s Hospital over the study period. Irrespective of the test used, there is an increase in Ct values with time after the first test. No participants with a hospital test Ct value below 26 produced a negative result within the short (12 day) study period, while several participants with a starting hospital Ct value above 26 were negative at subsequent time points.

**Figure 3.**
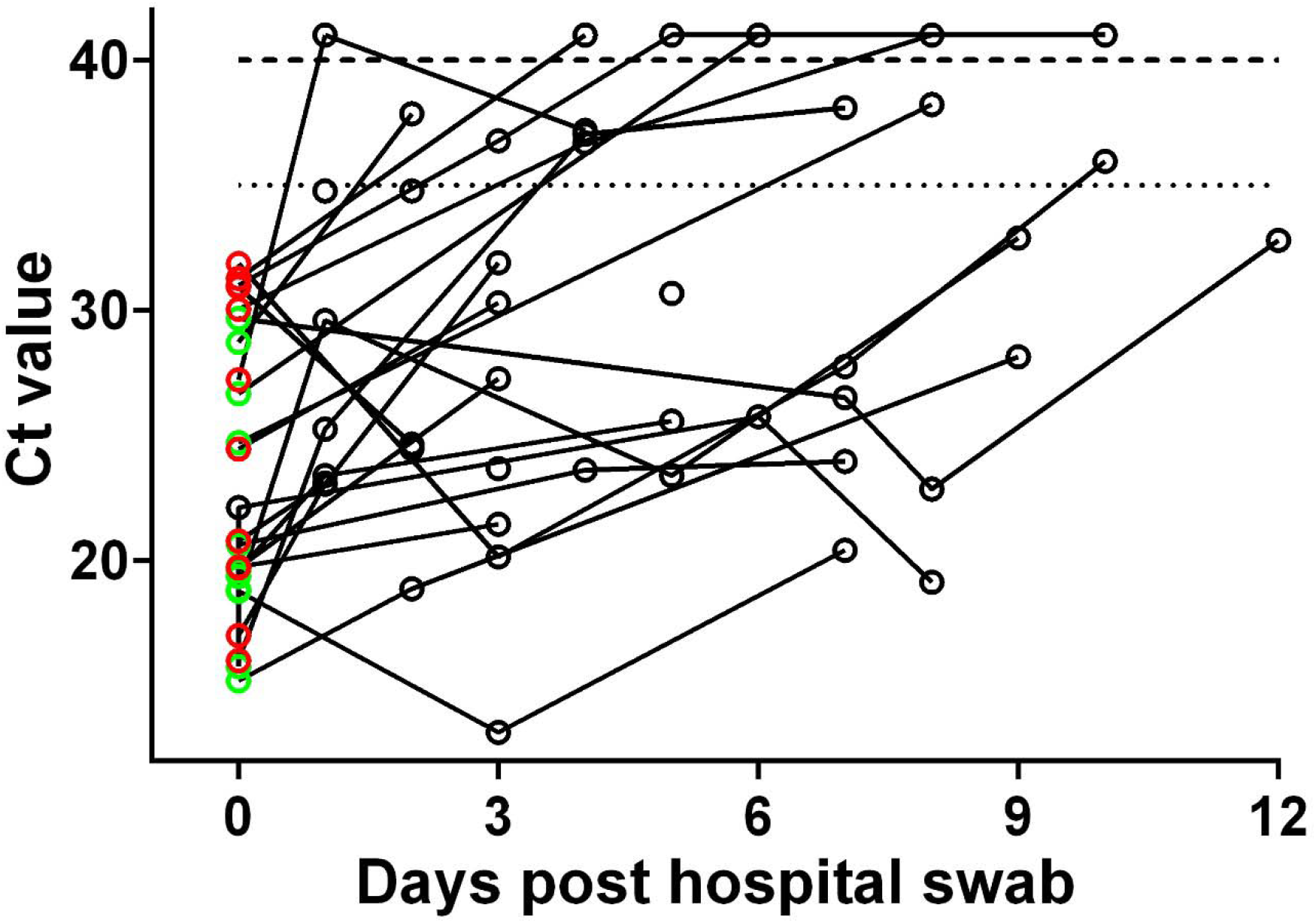
Longitudinal SARS-CoV-2 Ct values for COVID-19 patient samples. The hospital Ct results that were available are presented as the E gene Ct values for either the Roche (red symbols) or altona (green symbols) assays and used as day 0. Other data points are Ct values from the laboratory-based SAR-CoV-2 RT-PCR reference assay (black symbols). Any data points above the dashed line are negative and have nominally been given a Ct value of 41 to allow presentation. A Ct cut-off of 35 is represented by the dotted line. Note that both of the RT-PCR assays used by St George’s Hospital here have two independently measured targets, one of which is the E gene. The Ct values for the E gene target only were chosen to be presented since the average difference in Ct values between the E gene and the other target within each assay used were -0.18 ± 0.27 (mean ± SEM; n = 11; Range -2.2 to 0.45) and 0.47 ± 0.10 (mean ± SEM; n = 12; Range 0 to 1) for the Roche and altona assays, respectively.

Prior to the reference RT-PCR testing undertaken to derive the data presented in Figure 3, samples were run on the Q-POC™ SARS-CoV-2 RT-PCR assay. While the assay is designed to generate a SARS-CoV-2 positive or negative result, Ct values can also be derived. Figure 4 compares Ct values for the reference versus the Q-POC™ test. Data are from samples that were called positive by both tests (n = 32) and thus have paired Ct data. The two datasets passed normality tests (P = 0.86 and P = 0.69 for the reference and Q-POC™ tests, respectively; D’Agostino & Peatson omnibus normality test). Interestingly, there is very good concordance between the two paired datasets (as judged by eye, Fig.4A). Linear regression analysis of Q-POC™ and reference datasets (Fig. 4B) show a strong correlation (r^2^ = 0.76). A Bland-Altman plot demonstrated very good agreement between the 2 tests, with a bias of -6.7 Ct units meaning the Q-POC™ averages a 6.7 higher Ct value with mean (± SEM) Ct values of 25.8 ± 0.9 versus 32.5 ± 0.9 for the reference and Q-POC™ tests respectively. This measured bias is consistent with the Ct cut-off value of 40 (above which a result is called negative) for the reference assay compared with the Ct cut-off value of 45 for the Q-POC™ assay.

**Figure 4.**
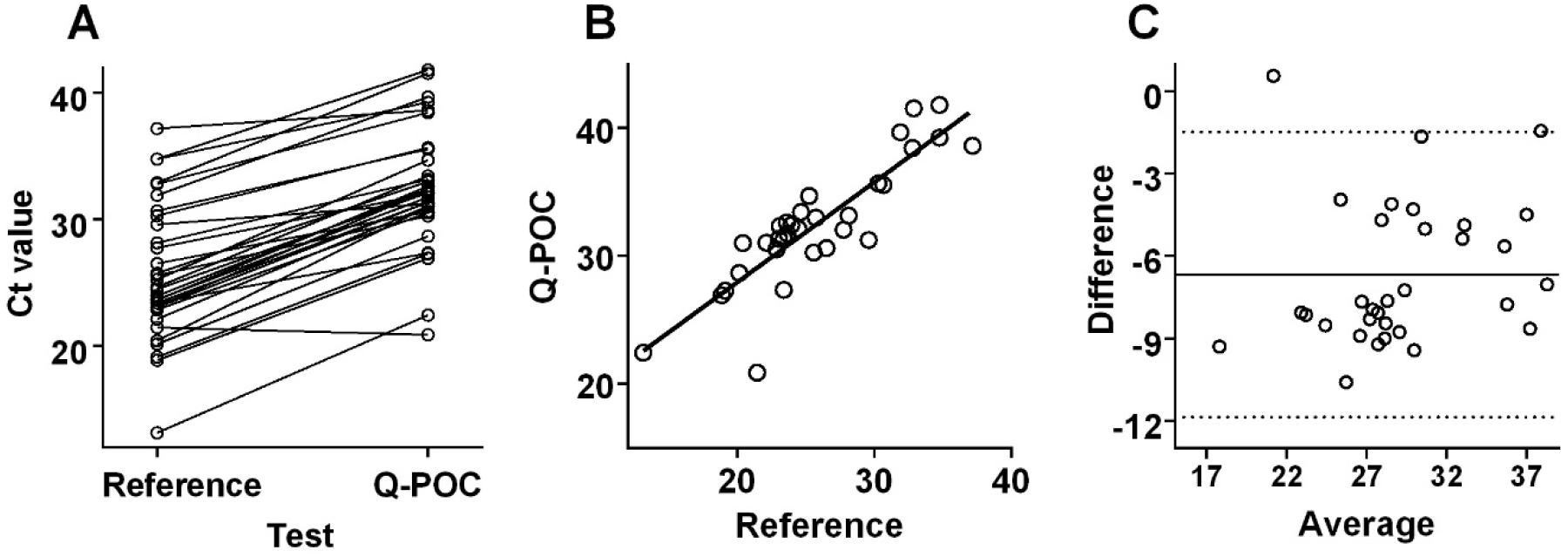
Comparison between Ct values generated by the laboratory-based SAR-CoV-2 RT-PCR reference assay and the Q-POC^™^ assay. A. Ct values for SARS-CoV-2 positive samples called by both the reference and Q-POC^™^ tests (n = 32) with Ct values below the 40 and 45 assay cut-off values, respectively. B. Linear regression plot to show the correlation between the reference and Q-POC^™^ assay Ct values. The line of best fit equation is y = 0.78x + 12.30 (r2 = 0.76). Bland-Altman plot to evaluate the agreement between the reference and Q-POC^™^ assay Ct values. The bias between the reference and the Q-POC^™^ test is -6.7 Ct units (solid line) and the 95% limits of agreement are denoted by the dotted lines.

With the inclusion of data from the 47 samples from COVID-19 patients and from single samples from 100 healthy volunteers the performance characteristics of the Q-POC™ test were determined (Table 2). The overall sensitivity (95% CI) of the Q-POC™ test was 80.0% (64.4-91.0%) against the SARS-CoV-2 RT-PCR laboratory reference test with a specificity and 99.1% (94.9-100%), with PPV and NPV of 97.0% and 93.0%, respectively. However, if the reference test Ct cut-off was adjusted to <35, sensitivity increased significantly to 96.9% (83.8-99.9%), while specificity dropped slightly to 98.3% (82.2-99.9%).

**Table 2.** Q-POC™ test performance.

| Cut-off (Ct) | Total | TP | TN | FP | FN | Sensitivity (95% CI) | Specificity (95% CI) | NPV (95% CI) | PPV (95% CI) |
| --- | --- | --- | --- | --- | --- | --- | --- | --- | --- |
| < 40 | 147 | 32 | 106 | 1 | 8 | 80.0%<br>(64.4-91.0) | 99.1%<br>(94.9-100) | 93.0%<br>(87.7-96.1) | 97.0%<br>(81.9-99.6) |
| < 35 | 147 | 31 | 113 | 2 | 1 | 96.9%<br>(83.8-99.9) | 98.3%<br>(93.9-99.8) | 99.1%<br>(94.3-99.9) | 93.9%<br>(79.7-98.4) |

Abbreviations: TP, true positives; TN, true negatives; FP, false positives; FN, false negatives; PPV, positive predictive value; NPV, negative predictive value; CI, confidence interval

Of note, while not evaluated in this study, the instructions for use (IFU; Supplementary material) for the Q-POC™ test states that the limit of detection is 1,000 genome equivalent copies per ml with a standard deviation of 0.8 and average Ct of 41.89, which is in line with the results presented above. Furthermore, *in vitro* analysis for exclusivity or cross-reaction was undertaken with no cross-reactivity observed under the tested conditions. All micro-organisms were used at a concentration of 1×10^5^ colony forming units (cfu) per reaction. Finally, as recommended by the Medicines & Healthcare products Regulatory Agency (MRHA), UK, QuantuMDx continuously monitor and assess the assay design *in silico* for newly emerging variants of SARS-CoV-2 with regular reports which are published by MHRA.

## Discussion

In a pandemic that causes a Public Health Emergency of International Concern, one of the earliest interventions is accurate diagnosis of an infection. At the start of a pandemic, this will depend largely on clinical evaluations (including imaging, biochemical and haematological assessments) that guide immediate containment measures. NAATs can be developed with urgency to diagnose a highly transmissible infectious disease accurately and assist in managing patients and breaking transmission chains (as happened with COVID-19 (2)). Accurate diagnostics are also critical to support the development of new therapeutic interventions, and to assess public health measures to contain infections, including the development and implementation of vaccination programmes.

There are several challenges for developers of diagnostics in the early stages of a pandemic. Regulatory authorities will be as unfamiliar with a disease such as COVID-19 as are other experts and this may delay appropriate evaluation of tests under development. Urgent need coupled with limited supplies and a motive to profit can encourage the proliferation of diagnostic tests that are easy to produce in bulk (such as lateral flow assays) but that fail to meet minimum requirements for accuracy (even if these requirements have been agreed, which itself can take too long procedurally). One way to circumvent some of these limitations is to repurpose high quality NAAT based testing platforms to deal with new infections such as caused by SARS-CoV-2.

The COVID-19 pandemic highlighted an urgent need for PoC methods to detect SARS-CoV-2, including its variants. To address this, QuantuMDx initially developed a laboratory based multiplex assay with simple to use lyophilised reagents that are stable when stored at room temperature, and quicker to run than many others (see (3)). However, these tests require automated equipment and batch testing, and cannot give rapid results for individual assessment.

QuantuMDx therefore repurposed cassettes, the portable analytical platform and software to provide an accurate, affordable and simple to use PoC test for SARS-CoV-2, as detailed in Methods. Here it is demonstrated that using a reference test Ct cut-of 35 that the Q-POC™ test for SARS-CoV-2 had a sensitivity of 96.88% (83.78%-99.92% CI) and a specificity of 98.3% (82.2-99.9%). These performance characteristics are comparable or better than other PoC NAAT devices (7-10).

Advantages of the Q-POC™ include a rapid run-time (∼32 minutes) compared with laboratory-based diagnostic platforms and some PoC NAAT devices also (e.g. (7, 8)). The Q-POC™ swab buffer is loaded directly into a fully sealed cartridge without the need for any extraction or sample processing steps. The sealed cartridge system also allows safe testing outside a laboratory, including primary care and community settings. Consistent with improvements in manufacturing, following preliminary evaluation (above) the cartridge failure rate (‘invalid result’ category) is expected to be < 2% and within specifications and industry standards. Centralised RT-PCR testing has the advantage of higher throughput capability with Q-POC™ being able to process only one cartridge at a time. This can be mitigated to an extent depending on the clinical setting and clinical testing algorithm by using multiple processing units with random access for each.

Ct values as an estimate of viral load and/or infectiousness can be influenced by many variables such as the site and methods for sample collection, the choice of diagnostic targets within the viral genome and sample quality (11). In establishing cut-off values a key principle is that the clinical risk of transmission must be minimised, so that chains of transmission are effectively interrupted. Several reports have suggested a Ct cut-off for reference RT-PCR tests of <35 (12-14). When Ct values are assessed in relation to the ability to culture virus results there are <3% culture positive samples with Ct = 35 (15). This Ct cut-off value is the most clinically relevant, because samples with higher cut-off Ct values are non-infectious and therefore could be termed ‘biological false positives’ despite being frequently (with up to 50% of positive results in assays having higher Ct cut-off values) detected (16). At a reference test cut-off Ct ≥35, the Q-POC™ test has excellent performance characteristics, that can be further examined in larger clinical trials. Testing strategies can be optimised for individual use cases, such as for travellers, health-care workers, prison services, triage of patients coming to emergency departments, wherever there is no laboratory and for contacts at risk of infection by timing testing strategies appropriately and by repeating testing at intervals when indicated.

Our study has several limitations. Although the sample size was small, it has been intended to assess and optimise performance of this novel technology not to develop individual use cases. Secondly, paired swab samples were not used for the reference and the Q-POC™ test rather both tests were run from a single swab sample. While this might remove the variable associated with multiple sampling from a participant subject at any given time point, the samples were placed into 3 ml of MSwab buffer (required for the Q-POC™ test) rather than the normal 1 ml of viral transport medium used in general for laboratory-based RT-PCR reference tests. This may have the effect of decreasing the reference test sensitivity by a small margin. Thirdly, the samples collected from the COVID-19 patients were not accordance with the IFU for the Q-POC™ test, which uses MN swabs. Further comparative studies would be needed to determine if nose and throat swabs can be used without loss of sensitivity, although it is worth noting that a small study found no difference in diagnostic sensitivity when comparing nose and throat and nasopharyngeal swabs (17).

## Supporting information

Supplemental Checklist

## Data Availability

All data referred to in the manuscript is present in the manuscript and supplementary material.

## Author contributions

Conceptualization: JC, MS, PS, CT, TP, HMS, SK. Methodology: JC, MS, KB, GM, PS, DEK, YA, CT, TP, HMS, SK. Investigation: KB, GM, DM, CP, A-MC, ES, JM, DJC, NE, EG, DEK, IM. Formal Analysis: JC, MS, DM, KB, YA, TP, HMS, SK. Writing – Original Draft: JC, YA, HMS, SK. Writing – Review & Editing: All.

## References

1. WHO. 2021. Coronavirus (COVID-19) Dashboard. https://covid19.who.int/. Accessed 24th June 2021.

2. Corman VM, Landt O, Kaiser M, Molenkamp R, Meijer A, Chu DK, Bleicker T, Brunink S, Schneider J, Schmidt ML, Mulders DG, Haagmans BL, van der Veer B, van den Brink S, Wijsman L, Goderski G, Romette JL, Ellis J, Zambon M, Peiris M, Goossens H, Reusken C, Koopmans MP, Drosten C. 2020. Detection of 2019 novel coronavirus (2019-nCoV) by real-time RT-PCR. Euro Surveill 25:2000045.

3. DHSC. 2020. QuantuMDx RT-PCR: SARS-CoV-2 Nucleic Acid Detection. https://assets.publishing.service.gov.uk/government/uploads/system/uploads/attachment_data/file/944909/QuantumDX_TVG_Report-V0.1-201208.pdf. Accessed 7th July 2021.

4. Cevik M, Tate M, Lloyd O, Maraolo AE, Schafers J, Ho A. 2021. SARS-CoV-2, SARS-CoV, and MERS-CoV viral load dynamics, duration of viral shedding, and infectiousness: a systematic review and meta-analysis. Lancet Microbe 2:e13–e22.

5. WHO. 2020. COVID 19 Public Health Emergencyof International Concern (PHEIC) - Global research andinnovation forum: towards a research roadmap. https://www.who.int/blueprint/priority-diseases/key-action/Global_Research_Forum_FINAL_VERSION_for_web_14_feb_2020.pdf. Accessed 7th July 2021.

6. Dinnes J, Deeks JJ, Adriano A, Berhane S, Davenport C, Dittrich S, Emperador D, Takwoingi Y, Cunningham J, Beese S, Dretzke J, Ferrante di Ruffano L, Harris IM, Price MJ, Taylor-Phillips S, Hooft L, Leeflang MM, Spijker R, Van den Bruel A, Cochrane C-DTAG. 2020. Rapid, point-of-care antigen and molecular-based tests for diagnosis of SARS-CoV-2 infection. Cochrane Database Syst Rev 8:CD013705.

7. Assennato SM, Ritchie AV, Nadala C, Goel N, Tie C, Nadala LM, Zhang H, Datir R, Gupta RK, Curran MD, Lee HH. 2020. Performance Evaluation of the SAMBA II SARS-CoV-2 Test for Point-of-Care Detection of SARS-CoV-2. J Clin Microbiol 59:e01262–20.

8. Gibani MM, Toumazou C, Sohbati M, Sahoo R, Karvela M, Hon TK, De Mateo S, Burdett A, Leung KYF, Barnett J, Orbeladze A, Luan S, Pournias S, Sun J, Flower B, Bedzo-Nutakor J, Amran M, Quinlan R, Skolimowska K, Herrera C, Rowan A, Badhan A, Klaber R, Davies G, Muir D, Randell P, Crook D, Taylor GP, Barclay W, Mughal N, Moore LSP, Jeffery K, Cooke GS. 2020. Assessing a novel, lab-free, point-of-care test for SARS-CoV-2 (CovidNudge): a diagnostic accuracy study. Lancet Microbe 1:e300–e307.

9. Hansen G, Marino J, Wang ZX, Beavis KG, Rodrigo J, Labog K, Westblade LF, Jin R, Love N, Ding K, Garg S, Huang A, Sickler J, Tran NK. 2021. Clinical Performance of the Point-of-Care cobas Liat for Detection of SARS-CoV-2 in 20 Minutes: a Multicenter Study. J Clin Microbiol 59:e02811–20.

10. Zhen W, Smith E, Manji R, Schron D, Berry GJ. 2020. Clinical Evaluation of Three Sample-to-Answer Platforms for Detection of SARS-CoV-2. J Clin Microbiol 58:e00783–20.

11. Rabaan AA, Tirupathi R, Sule AA, Aldali J, Mutair AA, Alhumaid S, Muzaheed, Gupta N, Koritala T, Adhikari R, Bilal M, Dhawan M, Tiwari R, Mitra S, Emran TB, Dhama K. 2021. Viral Dynamics and Real-Time RT-PCR Ct Values Correlation with Disease Severity in COVID-19. Diagnostics (Basel) 11:1091.

12. Chang MC, Hur J, Park D. 2020. Interpreting the COVID-19 Test Results: A Guide for Physiatrists. Am J Phys Med Rehabil 99:583–585.

13. Chen CJ, Hsieh LL, Lin SK, Wang CF, Huang YH, Lin SY, Lu PL. 2020. Optimization of the CDC Protocol of Molecular Diagnosis of COVID-19 for Timely Diagnosis. Diagnostics (Basel) 10:333.

14. La Scola B, Le Bideau M, Andreani J, Hoang VT, Grimaldier C, Colson P, Gautret P, Raoult D. 2020. Viral RNA load as determined by cell culture as a management tool for discharge of SARS-CoV-2 patients from infectious disease wards. Eur J Clin Microbiol Infect Dis 39:1059–1061.

15. Jaafar R, Aherfi S, Wurtz N, Grimaldier C, Van Hoang T, Colson P, Raoult D, La Scola B. 2021. Correlation Between 3790 Quantitative Polymerase Chain Reaction-Positives Samples and Positive Cell Cultures, Including 1941 Severe Acute Respiratory Syndrome Coronavirus 2 Isolates. Clin Infect Dis 72:e921.

16. Yang Q, Saldi TK, Gonzales PK, Lasda E, Decker CJ, Tat KL, Fink MR, Hager CR, Davis JC, Ozeroff CD, Muhlrad D, Clark SK, Fattor WT, Meyerson NR, Paige CL, Gilchrist AR, Barbachano-Guerrero A, Worden-Sapper ER, Wu SS, Brisson GR, McQueen MB, Dowell RD, Leinwand L, Parker R, Sawyer SL. 2021. Just 2% of SARS-CoV-2-positive individuals carry 90% of the virus circulating in communities. Proc Natl Acad Sci USA 118:e2104547118.

17. Vlek ALM, Wesselius TS, Achterberg R, Thijsen SFT. 2021. Combined throat/nasal swab sampling for SARS-CoV-2 is equivalent to nasopharyngeal sampling. Eur J Clin Microbiol Infect Dis 40:193–195.

