## Supplemental Material for "The QuantuMDx Q-POC™ SARS-CoV-2 RT-PCR assay for rapid detection of COVID-19 at point-of-care: preliminary evaluation of a novel technology"

### Q-POC™ SARS-CoV-2 Assay

Instructions for Use
Version Issued July 2021

For use with the QuantuMDx Q-POC™ point-of-care instrument

Reference Number: Q2700X

For *in vitro* Diagnostic Use Only

CE IVD

### 1. INTENDED USE

The Q-POC™ Severe Acute Respiratory Syndrome Virus Coronavirus-2 (SARS-CoV-2) Assay is a qualitative, real-time reverse transcription polymerase chain reaction (rtRT-PCR) assay for the detection of SARS-CoV-2 genomic RNA from upper respiratory specimens (e.g. nasal mid-turbinate swabs) collected from individuals who meet criteria for SARS-CoV-2 testing.
Results are used for the presumptive detection and identification of SARS-CoV-2 targeted genomic RNA sequences. SARS-CoV-2 genomic RNA is detectable in upper respiratory specimens during the acute phase of the infection and for a time after symptoms have abated. Positive results are indicative of active infection with SARS-CoV-2 but do not rule out co-infections with other viruses or bacteria. Clinical correlation with patient history and other diagnostic information is necessary to determine an individual's infection status. The detection of SARS-CoV-2 genomic RNA may not indicate the definitive cause of disease.
Negative results do not preclude SARS-CoV-2 infection and should not be used as a sole basis for treatment or other care management decisions. Negative results must be combined with clinical observations, the individual's history and epidemiological data and information.
The Q-POC™ SARS-CoV-2 Assay is intended for use by trained individuals.

### 2. PRINCIPLES OF THE PROCEDURE

The Q-POC™ SARS-CoV-2 Assay is a qualitative assay. The Q-POC™ SARS-CoV-2 Assay test cassette contains lyophilised reagents comprised of all components required for rtRT-PCR amplification and detection of genomic RNA from SARS-CoV-2 virus, a Specimen Process Control (SPC) and a reagent Rehydration Control (RC).

This assay is composed of two principal steps which are conducted automatically by the Q-POC™ instrument through manipulation of the Q-POC™ test cassette:

- Release of nucleic acids from the biological material contained in a sample under investigation, and;
- rtRT-PCR using oligonucleotide primers for the amplification of targeted genomic sequences and fluorogenic, hydrolysable DNA hybridisation probes for the specific detection of amplified target sequences.

This assay utilises RNase P as a SPC.
The assay utilises the fluorescent dye ROX as the RC.

The Q-POC™ instrument performs initial heat-based release of nucleic acids and subsequent reverse transcription of the SARS-CoV-2 genomic RNA and RNase P to synthesise complementary DNA (cDNA). Subsequently, the cDNA is amplified into specific DNA amplicons by real-time PCR.
The Q-POC™ SARS-CoV-2 Assay test cassette contains the primers and probes required for amplification of the SARS-CoV-2 and RNase P amplicons as detailed below.

| Analyte | Gene Targeted | Probe Fluorophore |
|---|---|---|
| SARS-CoV-2 | Orf1ab | FAM |
| SARS-CoV-2 | N | FAM |
| SARS-CoV-2 | S | FAM |
| Human Cells | RNase P | HEX |

### 3. Q-POC™ TEST CASSETTE

The Q-POC™ test cassette is a single-use consumable exclusively designed for use with the Q-POC™ point-of-care instrument. The test cassette is fully self-contained, including all reagents required to perform sample preparation, nucleic acid amplification and detection of SARS-CoV-2 RNA genomic targets and SPC by rtRT-PCR. Each test cassette comes individually packaged with a desiccant packet and the sample inlet cap attached to the cassette body for easy retrieval. Important features of the test cassette are indicated in the figure below.

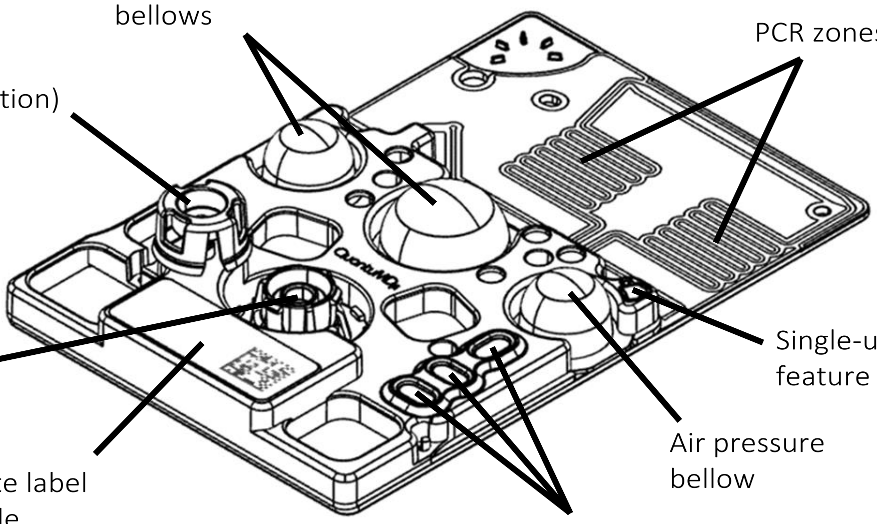

ⓘ Do not use any Q-POC™ test cassette if there are visible signs of damage to the test cassette or packaging.

ⓘ Do not use any test cassette which has the single-use feature depressed, as this cassette has previously been run in a Q-POC™ instrument and should be disposed of in an appropriate waste receptacle.

ⓘ Do not use any test cassette that has passed the expiration date marked on the cassette label or foil pouch label.

#### 4. ASSAY MATERIALS PROVIDED

**Q-POC™ SARS-CoV-2 Assay (Reference Number: Q2700X)**

| Kit Components | Kit Quantity |
|---|---|
| Q-POC™ SARS-CoV-2 Assay single-use test cassettes, individually sealed in a foil pouch | 120 test cassettes |

ⓘ Safety Data Sheets (SDS) are available upon request and on the QuantuMDx website at www.quantumdx.com.

#### 5. ADDITIONAL MATERIALS AVAILABLE FROM QUANTUMDX

**Q-POC™ SARS-CoV-2 Quality Control Kit (Reference Number: Q04-001-R01/ Q04-002-R02)**

| **Kit Components** |
|---|
| Q-POC™ SARS-CoV-2 Positive Control (Reference Number: Q04-001-R01) |
| Q-POC™ SARS-CoV-2 Negative Control (Reference Number: Q04-002-R02) |

**Pipette tips (Reference Number: Q15-004-P01/ Q15-003-P01)**

| |
|---|
| Aerosol-barrier, nuclease-free pipette tips, 20 µL (Reference Number: Q15-004-P01) |
| Aerosol-barrier, nuclease-free pipette tips, 1 mL (Reference Number: Q15-003-P01) |

**Q-POC™ 3mL Collection Kit (Reference Number Q14-116-P02)**

| **Kit Components** |
|---|
| Copan flexible FLOQSwabs®, individually wrapped (Reference Number: Q15-114-P01) |
| Specimen collection tube containing 3 mL of MSwab™ (Reference Number: Q03-112-R12) |

ⓘ Safety Data Sheets (SDS) are available upon request and on the QuantuMDx website at www.quantumdx.com.

#### 6. STORAGE AND HANDLING CONDITIONS

Q-POC™ SARS-CoV-2 Assay is shipped at ambient temperature.
Store Q-POC™ SARS-CoV-2 Assay test cassettes at ambient temperature in their original packaging. Once removed from its packaging, use the test cassette immediately.
Store Q-POC™ 3 mL Collection Kit and Q-POC™ SARS-CoV-2 Positive and Negative control vials as described in the accompanying Instructions for Use.

#### 7. EQUIPMENT AND DISPOSABLES REQUIRED

The following equipment and disposables are available from QuantuMDx:

- Q-POC™ point of care instrument
- Fixed volume pipette, 400 µL
- Fixed volume pipette, 20 µL
- 3 mL MSwab™ Collection Kit
- Aerosol resistant pipette tips
- Positive and Negative controls
- Printer
- USB key

The following equipment and disposables are required but NOT available from QuantuMDx:

- 2 to 8 °C refrigerator – to store the Q-POC™ SARS-CoV-2 Positive and Negative control vials.
- Personal Protective Equipment (PPE) – in accordance with local and institutional guidelines
- Biohazard waste facilities – in accordance with local and institutional guidelines

#### 8. FACILITY AND TRAINING REQUIREMENTS

Testing for the presence of SARS-CoV-2 genomic RNA should be performed in an appropriately equipped and maintained facility. Staff should be trained in the relevant technical and safety procedures.

ⓘ Please refer to the Center for Disease Control and Prevention (CDC) guidelines:
https://www.cdc.gov/coronavirus/2019-nCoV/lab-biosafety-guidelines.html

#### 9. PRECAUTIONS AND HANDLING REQUIREMENTS

##### Warnings and Precautions

- As with all testing and testing procedures, good laboratory practice is essential to ensure proper performance of the assay.
- For *in vitro* diagnostics (IVD) use.
- Positive results are indicative of the presence of SARS-CoV-2 RNA.
- Negative results should be treated as presumptive and, if inconsistent with clinical signs and symptoms or necessary for patient management, should be tested with an alternative molecular assay.
- The performance of this product has been tested only on nasal mid-turbinate specimen types.
- Appropriate PPE including lab coats, gowns, gloves, eye protection are recommended for manipulation of clinical specimens. All specimen processing should be performed in accordance with national, local and institutional biological safety recommendations.
- All specimens and samples should be handled as infectious, using good laboratory procedures and Universal Precaution. Only trained health professionals should perform this procedure.
- Positive and negative control material includes substances of human and animal origin, attention must be drawn to their potentially infectious nature.
- If spillage should occur on the cassette or workspace, immediately disinfect with a freshly prepared 0.5% solution consisting of household bleach (5-10% sodium hypochlorite) diluted in distilled or deionized water or follow appropriate procedures as outlined by the site or facility.
- Consult the Q-POC™ instrument User Manual for instructions on cleaning procedures for the exterior of the instrument.
- Never clean the interior surfaces of the Q-POC™ instrument, unless instructed otherwise by QuantuMDx or an authorised representative.

- Closely follow procedures and guidelines provided to ensure that the assay is performed correctly. Any deviation from the procedure and guidelines may affect optimal performance of the assay.
- False positive results may occur if carryover is not adequately controlled during specimen and sample handling and processing.

##### Specimen and Test Cassette Handling

- Wear appropriate powder-free gloves when handling specimens and reagents.
- It is advised to perform specimen collection, handling, and test cassette preparation procedures in a well-ventilated area following local risk assessment and PPE requirements, as necessary.
- Wash hands thoroughly after handling specimens, samples, kit reagents and after removing one's gloves.
- Use the sterile aerosol-barrier, nuclease-free pipette tips provided when handling specimens, samples, and reagents for the detection of RNA.
- Handle all reagents, controls, specimens and samples according to good laboratory practice in order to prevent carryover and/or contamination of specimens, samples or controls.
- Before use, visually inspect the collection kit to ensure that there are no signs of breakage or leakage. If there are signs of breakage or leakage, do not use that material for testing and contact QuantuMDx Customer Support immediately.
- Before use, visually inspect the Q-POC™ test cassette to ensure that there are no signs of breakage or leakage. If there are signs of breakage or leakage, do not use that material for testing and contact QuantuMDx Customer Support immediately.
- Upon visual inspection of the cassette, ensure the single-use feature located on the top right corner of the teal plastic cassette cover is intact. Do not use if the feature is deformed, as this indicates the test cassette has already been used.
- Do not place any stickers on the barcode, or on any of the functional areas of the cassette.
- Do not expose the cassette to direct sunlight.
- Do not handle the cassette by the film.
- Do not eat, drink, smoke, apply cosmetics or handle contact lenses in areas where reagents, specimens and samples are handled.
- Decontaminate and dispose of all potentially infectious materials in accordance with institutional, local and relevant national regulations.
- Dispose of all cleaning materials as biological waste.
- Amplification technologies such as RT-PCR are sensitive to accidental introduction of product from previous amplification reactions. Incorrect results could occur if either the clinical specimen or the real-time reagents used in the amplification step become contaminated by accidental introduction of amplification product. Measures to reduce the risk of contamination in the laboratory include physically separating the activities involved in performing RT-PCR in compliance with good laboratory practices and establishing a unidirectional workflow. Since the Q-POC™ SARS-CoV-2 Assay single-use test cassette is a fully enclosed device, the risk of amplicon contamination is minimal.
- Avoid microbial and nuclease contamination of the specimen. Use the sterile, aerosol-barrier pipette tips available from QuantuMDx and change between all liquid transfers.
- Work area, instrumentation and equipment must be considered potential sources of contamination. Change gloves after contact with potential contaminants (specimens, eluates, and/or amplified product) before handling unopened reagents, controls or specimens.
- Check the collection kit expiry date before use.
- Do not use Q-POC™ test cassettes or other components beyond their recommended expiry dates.
- Set up each run separately.
- Dispose of unused test cassettes and human specimens according to all relevant local and national regulations.

#### 10. PREVENTION OF NUCLEIC ACID CONTAMINATION

The possibility of nucleic acid contamination is minimized when:

- Aerosol-barrier pipette tips are used for all pipetting. The pipette tips are discarded after use.
- Work surfaces, instrumentation and equipment are regularly cleaned with the appropriate solutions.
- Appropriate PPE is utilised, and good laboratory practices are followed for all procedures.

#### 11. SAMPLE COLLECTION, HANDLING AND STORAGE

**Collecting a Nasal mid-turbinate (NMT) Specimen:**

ⓘ Refer to the QuantuMDx Q-POC™ 3 mL Collection Kit (Reference Number Q15-001-P01) Instructions for Use or your facility's procedures for more details. Important features of the collection swab are reported below. DO NOT touch the swab applicator in the area beyond the moulded breakpoint to prevent contamination of the swab. Should the swab be handled anywhere beyond the breakpoint indicator, dispose of it in an appropriate waste bin and get a fresh swab package. Use of aseptic technique is recommended to avoid contamination.

**Collection Swab**

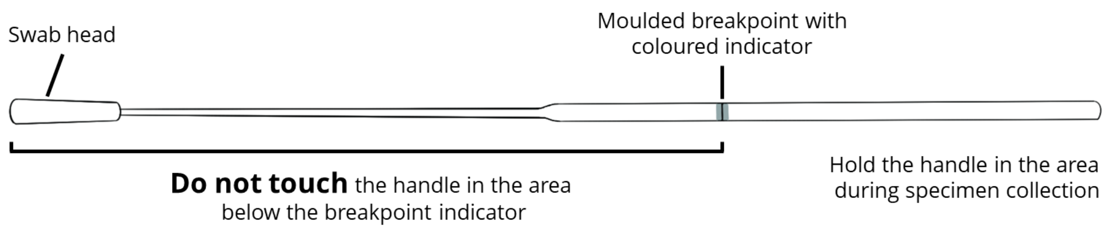

1. Open the kit package and remove the tube of MSwab™ reagent and the inner pouch containing the sterile swab applicator.

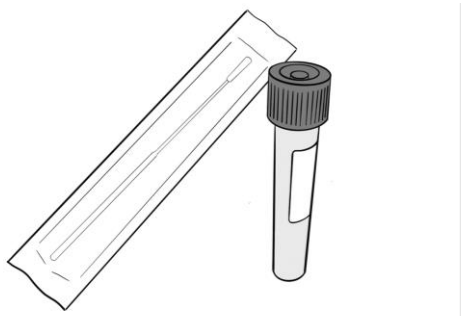

2. Remove the swab applicator from its peel pouch and use it to collect the clinical specimen.

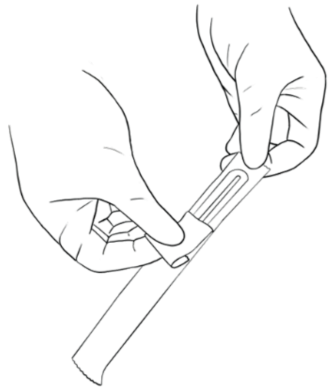

CONTINUED ON NEXT PAGE

3. Using gentle rotation and tilting patient's head back 70 degrees, insert the swab about 2 cm into the nostril without tipping the swab head up or down.

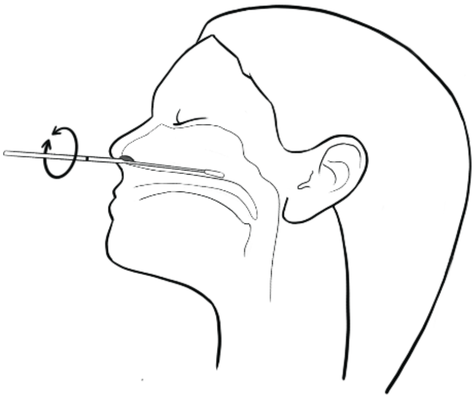

ⓘ The nasal passage runs parallel to the floor, not parallel to the bridge of the nose.

4. Leave the swab in place for a few seconds, then slowly rotate the swab as it is being withdrawn.

⚠ DO NOT USE FORCE while inserting the swab. It should travel smoothly with minimal resistance. If resistance is encountered, withdraw the swab a little bit, without taking it out of the nostril, then elevate the back of the swab and move it forward.

At all times when handling the swab applicator, the operator must NOT touch the area below the breakpoint line, as this may result in contamination.

5. Unscrew the cap of the MSwab™ tube and insert the swab until the marked breaking point.

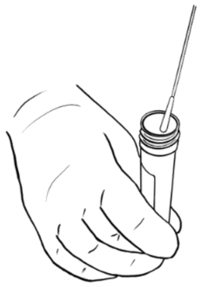

6. Bend and break the swab at the marked breaking point holding the tube away from your face.

Hold the tube away from your face while breaking the swab handle to prevent potential exposure to biological material.

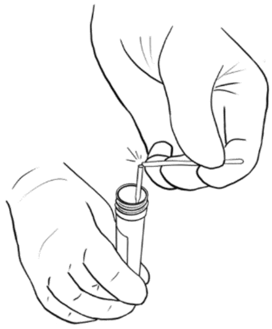

⚠ DO NOT place the swab on a bench. Use of aseptic technique is recommended to avoid contamination.

7. Discard the broken handle into an approved biohazard disposable container.

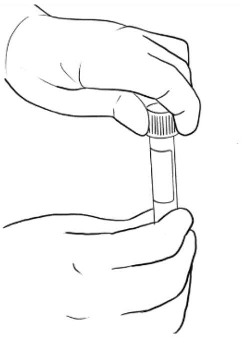

8. Replace cap on the tube and secure tight to prevent leaks.

9. Invert the tube five (5) times.

The specimen is now ready to be loaded onto the Q-POC™SARS-CoV-2 Assay test cassette.

#### 12. STORING SPECIMENS

Store specimens refrigerated (2 to 8 °C) for up to 24 hours prior to processing.

ⓘ IMPORTANT:

The following can affect the results obtained:

- Inadequate or inappropriate collection of specimens
- Incorrectly stored specimens and samples
- Incorrectly transported specimens
- Use of non-validated specimen types
- Inadequate specimen volume

#### 13. SAMPLE PREPARATION

The Q-POC™ SARS-CoV-2 Assay does not require external extracting or enriching of nucleic acids from specimens. The operator is only required to load the designated volume of specimen to the test cassette as described below.

#### 14. QUALITY CONTROL PREPARATION

Please consult the Q-POC™ SARS-CoV-2 Quality Control kit (Reference Number: Q04-001-R01/ Q04-002-R02) package insert for instructions on the preparation of the quality controls for use with the Q-POC™ SARS-CoV-2 Assay test cassettes.

SARS-CoV-2 Assay Controls:

Specimen Process Control (SPC) – the human RNase P gene is utilised by the assay to provide guidance as to the performance of specimen acquisition, performance of the nucleic acid release process, the performance of the assay chemistry and real-time instrumentation. The RNase P gene is naturally present in human cells and should be present in all correctly collected human specimens. The SPC has been designed not to compete with the detection of SARS-CoV-2 target loci. The SPC signal should be present in all negative samples and most SARS-CoV-2 positive samples. However, in the presence of high concentrations of SARS-CoV-2 genomic RNA the RNase P signal may not be detected. The reactivity of the SPC is part of the validity and acceptance criteria for every batch run of the assay.

Negative Control (NC) – NC should be run according to local procedures. The NC contains a standardised concentration of human cells to be a target for the human RNase P SPC detection reagents contained within the Q-POC™ SARS-CoV-2 Assay. The NC is needed to determine if carry-over contamination of SARS-CoV-2 has occurred during routine preparation of the test cassettes. The NC is supplied in the Q-POC™ SARS-CoV-2 Quality Control kit (Reference Number: Q04-002-R02).

Positive Control (PC) – PC should be run according to local procedures. The PC contains a standardised concentration of inactivated intact SARS-CoV-2 virus and human cells as targets for the SARS-CoV-2 and human RNase P SPC detection reagents contained within the Q-POC™ SARS-CoV-2 Assay, respectively. The PC is utilised for validity and acceptance criteria for both the assay chemistry and the Q-POC™ instrument. The PC is supplied in the Q-POC™ SARS-CoV-2 Quality Control kit (Reference Number: Q04-001-R01).

No Template Control (NTC) – It is recommended to periodically run an NTC comprised of MSwab™ reagent only (3 mL vial, Reference Number Q03-112-R12).

#### 15. ASSAY PROCEDURE

The QuantuMDx Q-POC™ SARS-CoV-2 Assay does not require enrichment of nucleic acids from the test specimen prior to loading onto the Q-POC™ test cassette.

Consult the User Manual for guidance on the proper operation of the Q-POC™ instrument.

Once the sample to be tested is added, use the test cassette immediately.

#### 16. PREPARING THE SPECIMEN AND TEST CASSETTE

**Handling of the test specimen and loading the Q-POC™ test cassette should be performed using personal protective equipment and should be performed in accordance with national, local and institutional biological safety recommendations. All biological samples, spills and leaks should be considered as biohazardous**.

1. Mix the contents of the tube thoroughly by completely inverting the tube five (5) times.

AVOID shaking the tube contents to prevent the formation of excess bubbles in the liquid solution.

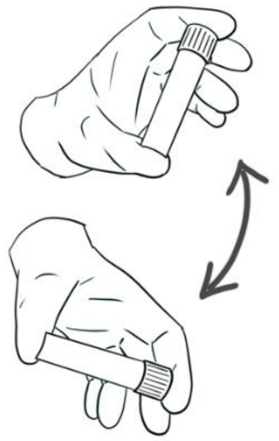

2. Remove the specimen tube cap.

3. In an aseptic manner, transfer 400 µL of the sample from the specimen collection tube into the test cassette sample inlet port using the fixed volume pipette and an aerosol barrier pipette tip. Discard used pipette tip into an approved biohazard disposable container.

AVOID introducing bubbles into the inlet port when pipetting the sample into the cassette.

When loading the test cassette, the pipette should be held at a 45° angle. Release the fluid in a steady motion, pointing the pipette tip in the direction of the centre of the cassette.

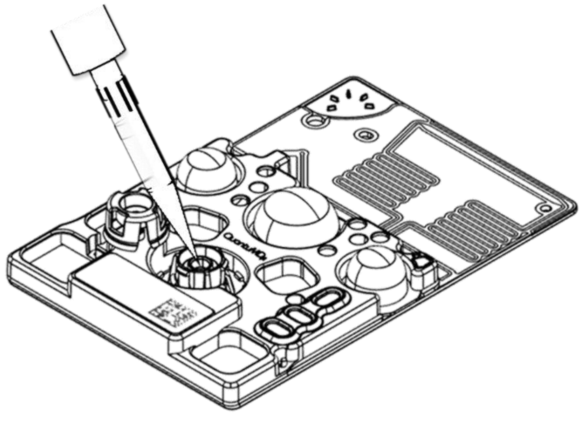

CONTINUED ON NEXT PAGE

4. Remove the cassette cap from the test cassette body and seal the sample inlet port by aligning the cap with the retention clips. Press firmly to ensure the cap is locked in place.

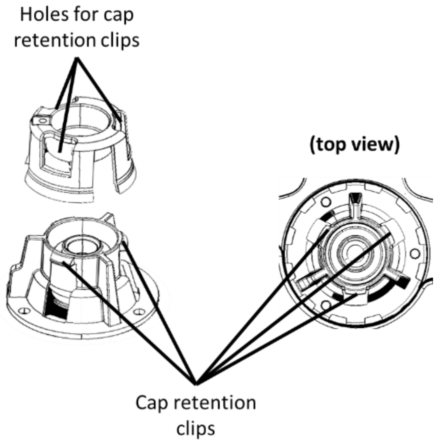

ⓘ A 'click' should be heard and/or felt when the cap is locked into place correctly.

⚠ **DO NOT** remove the cap once it has been placed.

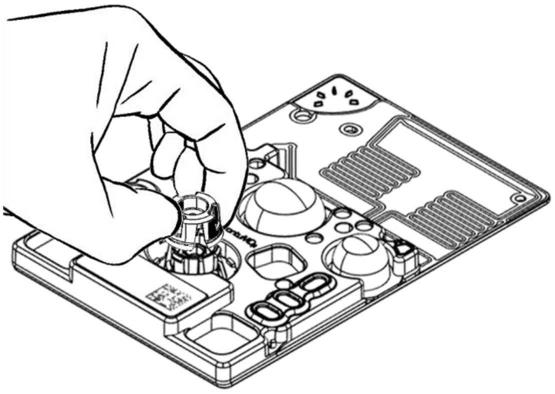

5. Using a permanent pen, write the sample ID on the cassette label using the blank space provided.

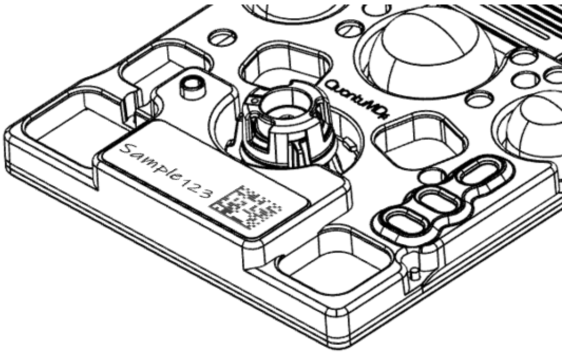

⚠ **DO NOT** place any labels on the test cassette, except in the blank space provided on the cassette label. Labels placed on other surfaces of the cassette may disrupt proper functioning.

⚠ **DO NOT** write on or obstruct the 2D barcode on the bottom-right side of the test cassette label.

The test cassette is now ready to load into the Q-POC™ for testing.

#### 17. Q-POC™ OPERATION

The Q-POC™ SARS-CoV-2 Assay test cassette is intended to be processed exclusively on the Q-POC™ point-of-care instrument.

1. Press the **RUN A TEST** button on the **HOME** screen.

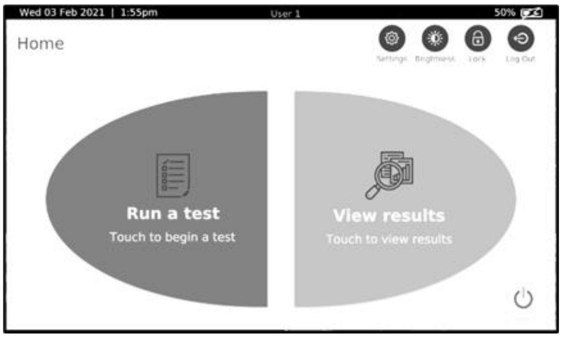

CONTINUED ON NEXT PAGE

2. Press **CLICK TO SCAN** button to activate the scanner. Place the cassette 2D barcode on the cassette label within the barcode reader alignment beam to help with the proper positioning. Barcodes should be placed in the proximity of the reader window to be successfully scanned.

ⓘ Use **ONLY** Q-POC™ SARS-CoV-2 Assay test cassettes with the Q-POC™ instrument.

ⓘ **DO NOT** use test cassettes that have passed the expiry date or previously used cassettes.

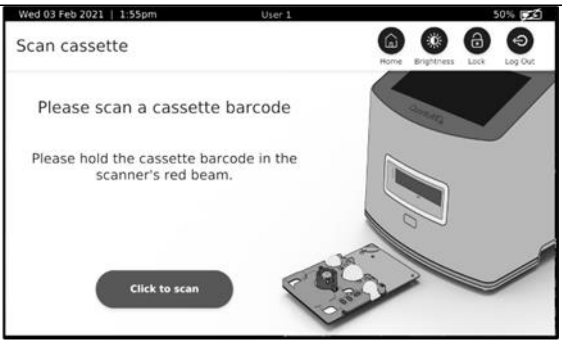

3. Type the Sample ID (e.g. sample123) into the blank data field.
4. Press the **NEXT** button.

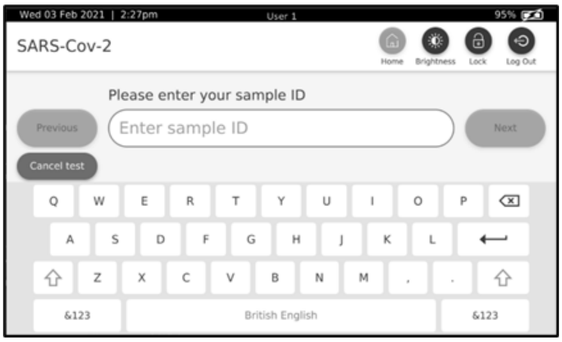

5. Type the patient age in years (e.g. 50) into the blank data field.
6. Press the **NEXT** button.

ⓘ This step is optional and may be skipped by touching the **SKIP** button.

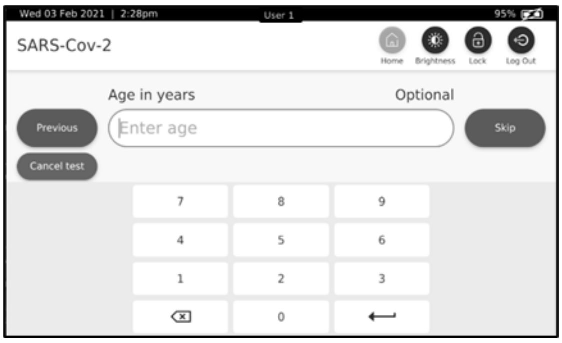

7. Press the button from the menu that corresponds to the patient's sex.
8. Press the **NEXT** button.

ⓘ This step is optional and may be skipped by touching the **SKIP** button.

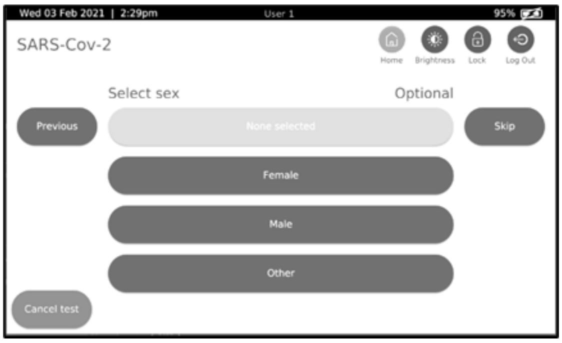

9. Press the empty "Test Notes" field.
10. Type any additional notes into the field.
11. To start a new line of text press the enter key ⟵ on keyboard.
12. Press **NEXT** button.

ⓘ This step is optional and may be skipped by touching the **SKIP** button.

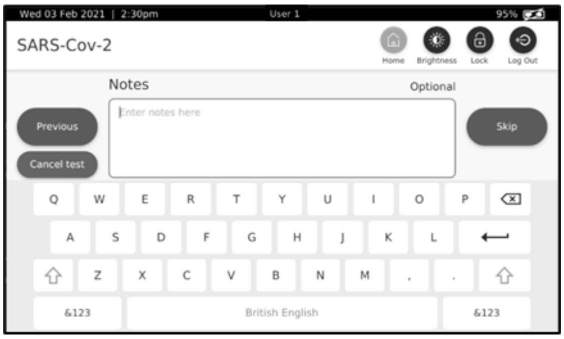

CONTINUED ON NEXT PAGE

13. Review the **TEST DETAILS SUMMARY** for accuracy and press **EDIT** to update any information, if required.

14. Press **CONFIRM** to proceed to the next step.

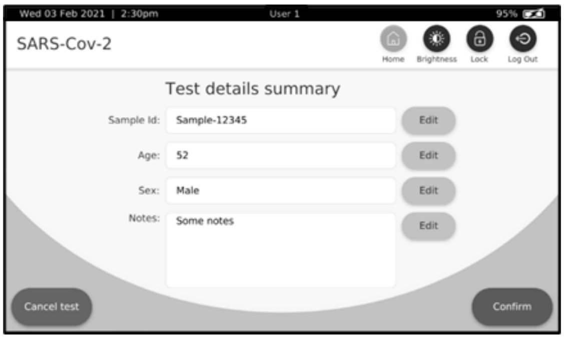

15. Press the **OPEN DOOR** button.

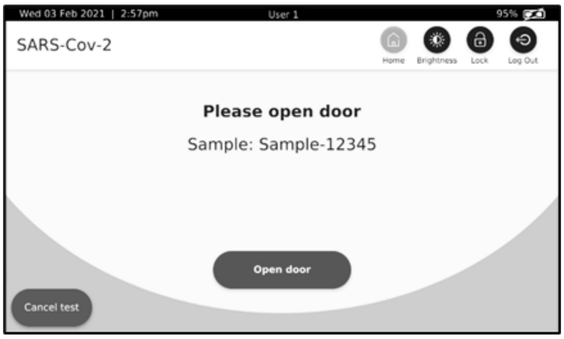

16. When prompted by the software, insert the test cassette into the cassette loading bay, as illustrated.

ⓘ The LED lights surrounding the cassette loading bay will begin to flash when the instrument is ready to accept the test cassette.

⚠ All biological samples, spills and leaks should be classed as biohazardous. Appropriate personal protection and handling procedures should be used.

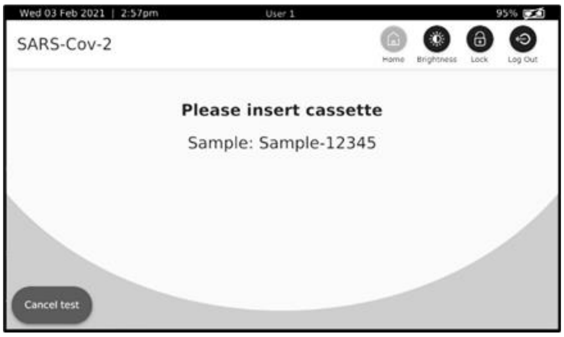

17. Place the thin, black portion of the test cassette flat on the bottom of the cassette loading bay and slide the test cassette into the instrument until it comes to a stop.
    When inserted, approximately three (3) cm of the test cassette will remain exposed until the instrument scans and draws the entire cassette into the loading bay and closes the door.

ⓘ **DO NOT** apply excessive force when inserting the cassette. The cassette should slide into the Q-POC™ easily.

⚠ **NEVER** place hands or fingers directly into the open cassette loading bay.

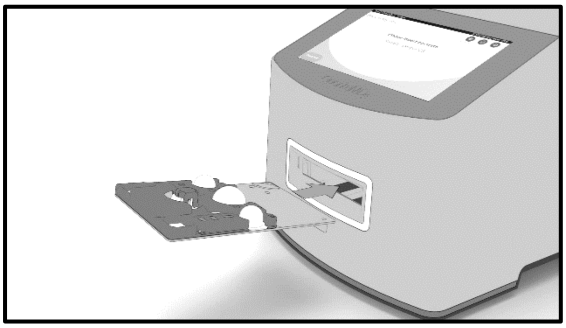

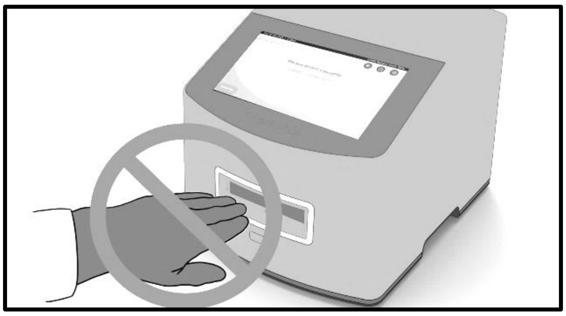

CONTINUED ON NEXT PAGE

18. During testing, the software will provide test details and the test run progress status on the display.

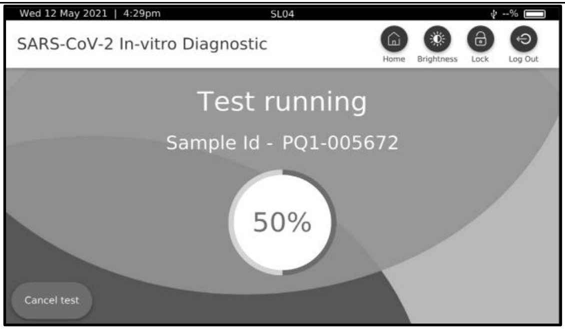

19. When the test has completed, press the EJECT CASSETTE button to open the cassette bay door and unload the used test cassette.

ⓘ If the cassette is too hot to eject, a warning message will appear. Allow approximately 30 seconds to sufficiently cool the used test cassette.

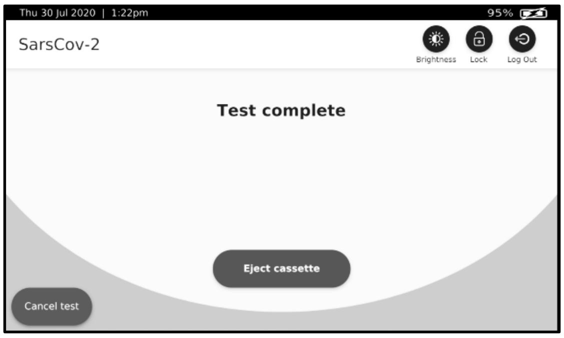

20. Remove the used test cassette and dispose of it in an appropriate waste container.

☣ All biological samples, spills and leaks should be considered as biohazardous and appropriate personal protection and handling procedures should be employed.
Test cassettes containing a specimen should be considered biohazardous and should be disposed of in an appropriate waste container, according to local, regional and national guidelines and regulations.

⚠ DO NOT attempt to open the used test cassette.

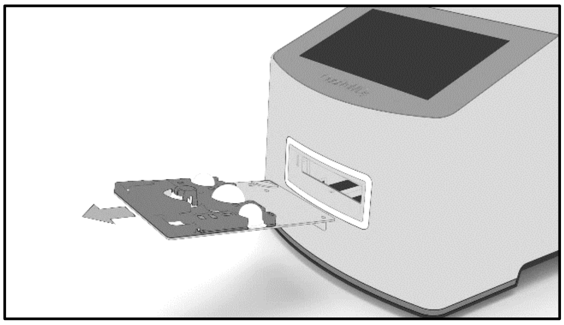

21. Review the test results after the run has completed.

22. Tap HOME to return to the Home screen.

ⓘ Press VIEW GRAPH to display the amplification curves or ALL RESULTS to view previous test results.
Press EXPORT to immediately upload the test result onto a USB storage drive. Refer to the Q-POC™ User Manual for more details.

⚠ See the Interpretation of Results section of this document for more details on reporting of results. DO NOT use amplification curves for diagnostic purposes, as these are provided for informational use ONLY. Please always refer to the "Result" screen for result interpretation.

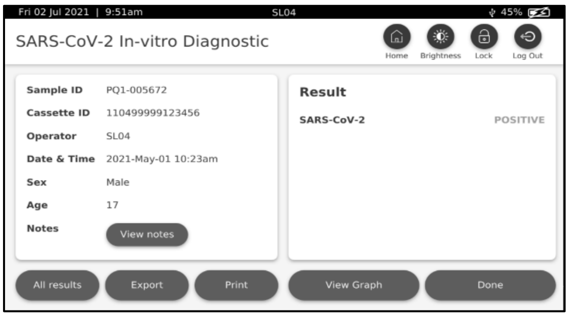

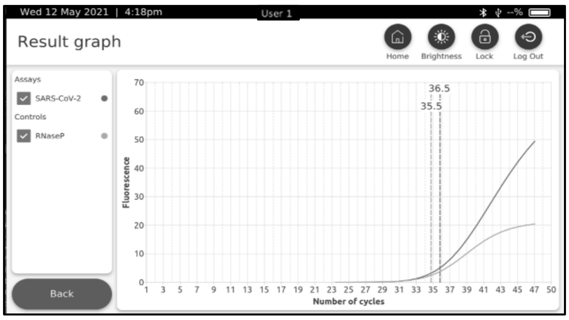

END

#### 18. ASSAY SPECIFIC QUALITY CONTROLS

The Q-POC™ SARS-CoV-2 Assay includes quality controls for monitoring the performance of the test during each test run. These quality controls are described below.

| Control Type | Used to Monitor |
|---|---|
| SPC | Sample extraction efficiency<br>PCR inhibition<br>Process error |
| RC | Correct rehydration of lyophilised reagents in the test cassette |

Sample Process Control (SPC) – The human RNase P gene is utilised in the assay to provide information about the performance of: specimen acquisition, the extraction/enrichment process, the assay reagents and the real-time instrumentation. The SPC should be present in all correctly collected human specimens. The SPC has been designed not to compete with the detection of SARS-CoV-2 targeted loci. The SPC signal should be present in all negative samples and most SARS-CoV-2 positive samples. However, in the presence of high concentrations of SARS-CoV-2 genomic RNA, the RNase P signal may not be detected because of strong competition for PCR reagents from the SARS-CoV-2 reaction. This sample result is still valid and acceptable. The reactivity of the SPC is part of the validity and acceptance criteria for every batch run of the assay.
Failure of an SPC for a sample only invalidates the sample result when the sample is expected to have the presence of human RNase P and is negative for the presence of SARS-CoV-2.

Rehydration Control (RC) – The rehydration control is the fluorescence dye ROX, which is contained within the lyophilised reagents in the test cassette. Detection of ROX is part of the validity criteria for a run. The validity of a test run is determined automatically by the Q-POC™ using a software algorithm to analyse various assay parameters.

In the event of a higher-than-expected numbers of test failures and/or suspected inaccurate test results, please contact QuantuMDx Customer Support for assistance.

#### 19. EXOGENOUS ASSAY QUALITY CONTROLS

Quality control requirements should be performed in conformance with local and national regulations or accreditation requirements and your laboratory's standard quality control procedures. Quality control procedures are intended to monitor reagent and assay performance.

| Control Type | Used to Monitor |
|---|---|
| Positive (PC) | Substantial reverse transcriptase and polymerase failure<br>Substantial primer and probe failure |
| Negative (NC) | Reagent contamination<br>Environmental contamination |

Exogenous assay quality controls are provided with the test and additional controls may be purchased separately. Positive and Negative controls are provided as 6 x 500 µL vials in each control kit, including the following:

| Material | Virus / Cell line | Kit Contents | Reference Number |
|---|---|---|---|
| Positive Control | SARS-CoV-2 (Isolate: USA-WA1/2020)/ Human A-549 cells | 6 vials, 0.5 mL each | Q04-001-R01 |
| Negative Control | Human A-549 cells | 6 vials, 0.5 mL each | Q04-002-R02 |

Exogenous assay quality controls should be prepared and tested following local procedures to ensure proper functioning of the Q-POC™ instrument and the Q-POC™ SARS-CoV-2 Assay. It is recommended to test each new assay lot with exogenous positive and negative controls before commencing the testing of specimens.
Failure of the controls (PC or NC) invalidates the run. Results should not be reported as repeat testing should

be done starting from original specimen, using a new aliquot of positive control. If repeat results are still invalid, results should not be reported, and testing should be repeated from the original specimen, or a new specimen should be collected and tested.
Failure of an SPC for a sample invalidates the sample result when the sample is negative for the presence of SARS-CoV-2. In most cases the SPC signal should be present for a SARS-CoV-2 positive sample, however, the presence of high concentrations of SARS-CoV-2 genomic RNA the SPC signal may not present. This sample result is still valid and acceptable.
In the case of higher-than-expected numbers of test failures and/or suspected inaccurate test results, please contact QuantuMDx Customer Support.

#### 20. INTERPRETATION OF RESULTS

⚠ **ONLY report test results as displayed by the test result screen of the Q-POC™. The amplification graph is only provided for informational purposes and to allow visualisation of the amplification curves.**

The Q-POC™ instrument automatically analyses and interprets test results using the criteria described below. All instances of SARS-CoV-2 amplification of Ct ≤ 45 cycles indicate a SARS-CoV-2 positive result.

| SARS-CoV-2 (FAM) | RNase P (HEX) | Interpretation |
|---|---|---|
| + | + | Positive for the presence of SARS-Cov-2 genomic RNA<br>Valid run |
| + | -* | Positive for the presence of SARS-Cov-2 genomic RNA<br>* in presence of high concentrations of SARS-CoV-2 genomic RNA the SPC signal may not present<br>Valid run |
| - | + | Negative for the presence of SARS-CoV-2 genomic RNA<br>Valid run |
| - | - | Invalid run; no specimen detected, determine root cause and take appropriate action as outlined below |

Additionally, for the Q-POC™ to display a valid test result, all assay specific quality controls (e.g. SPC and RC) and other system checks must be valid and acceptable. Should any of these quality checks not pass, the Q-POC™ will report the test result as invalid. It is recommended to retest invalid test sample using a new test cassette.
If the retest result remains invalid due to non-amplification of the SPC (when known human cells are present), this may indicate the presence of PCR inhibitors in the sample. A new sample should be obtained and tested. The results should be reported as 'Void Test Result due to Inhibition' if a sample repeatedly shows no amplification of the SPC when known human cells are present. However, in the presence of high concentrations of SARS-CoV-2 genomic RNA, the RNase P signal may not be detected because of strong competition for PCR reagents from the SARS-CoV-2 reaction. This sample result is still valid and acceptable.

#### 21. LIMITATIONS

Training is necessary prior to performing the assay.
Performance of the Q-POC™ SARS-CoV-2 Assay has only been established in nasal mid-turbinate swab specimens. Other specimen types may yield inaccurate results.
The detection of viral nucleic acid is dependent upon proper specimen collection, handling, transportation, storage, and preparation. Failure to observe proper procedures in any one of these steps can lead to incorrect results. There is a risk of false negatives resulting from improperly collected, transported, or handled specimens. False negative results may also occur in the presence of amplification inhibitors, or if inadequate numbers of organisms are present in a clinical sample. As for any molecular test, there is a risk of false negative results due to the presence of sequence variants in viral targets of the assay. QuantuMDx believes in vigilance and as such is regularly, *in silico*, re-evaluating the performance of its assay versus all newly deposited SARS-CoV-2 sequences, including emerging variants. Latest reports can be accessed by contacting QuantuMDx Customer Support.
This assay cannot rule out diseases caused by other bacterial or viral pathogens.
The prevalence of infection will affect the test's predictive value.
A trained health professional should interpret test results in conjunction with the Persons Under Investigation (PUI's) medical history, clinical signs and symptoms, and the results of other tests.
Analyte targets (viral nucleic acids) may persist *in vivo*, independent of virus viability. Detection of the analyte does not imply that the corresponding virus is infectious or is the causative agent for clinical symptoms.
A sample yielding negative results can contain pathogens other than SARS-CoV-2.

#### 22. PERFORMANCE CHARACTERISTICS

The Limit of Detection (LoD) or analytical sensitivity was determined as the lowest concentration of SARS-CoV-2 target, that could be detected by the Q-POC™ SARS-CoV-2 Assay with a ≥95% positivity rate.

| SARS-CoV-2 Copies/mL | Replicates | Average Ct | Standard Dev. | RNase P Copies/mL | Average Ct | Standard Dev. | Sensitivity |
|---|---|---|---|---|---|---|---|
| 1,000 | 5/5 | 41.89 | 0.80 | 500 | 34.11 | 0.32 | 100% |
| 800 | 4/5 | 42.51 | 2.42 | 500 | 33.99 | 0.97 | 80% |
| 600 | 4/5 | 43.00 | 0.76 | 500 | 33.94 | 0.47 | 80% |
| Negative | 0/5 | NA | NA | 500 | 34.08 | 1.11 | NA |

Presumptive LoD determined by amplification of different concentrations of SARS-CoV-2 RNA in the presence of a fixed concentration of RNase P.

The LoD was verified by running 21 replicates below, at and above the LoD determined above. The data is presented in the table below and demonstrates a sensitivity of ≥95% at the LoD (1,000 c/mL SARS-CoV-2).

| SARS-CoV-2 Copies/mL | Replicates | Average Ct | Standard Dev. | RNase P Copies/mL | Average Ct | Standard Dev. | Sensitivity |
|---|---|---|---|---|---|---|---|
| 1,200 | 21/21 | 40.65 | 1.75 | 500 | 34.47 | 0.52 | 100% |
| 1,000 | 20/21 | 41.89 | 1.64 | 500 | 35.57 | 1.25 | ≥95% |
| 800 | 19/21 | 42.69 | 1.37 | 500 | 35.09 | 0.83 | <95% |
| Negative | 0/10 | NA | NA | 500 | 35.20 | 1.75 | NA |

#### 23. INCLUSIVITY (REACTIVITY)

Sequences for the SARS-CoV-2 virus were downloaded from the NCBI and GISAID. Sequences were filtered to remove duplicate downloads, then filtered on size (≥ 25,000 bases in length) to ensure that only full or close full-length sequences were used for inclusivity testing. The downloading and vetting of the SARS-CoV-2 sequence data resulted in 10,151 sequences (1,022 = NCBI and 9,126 = GISAID) utilized for inclusivity analysis. Each primer and probe of the Q-POC™ SARS-CoV-2 Assay were then analyzed versus each SARS-CoV-2 sequence one at a time to ensure that they complemented the SARS-CoV-2 sequence at the desired position. Mismatching was then checked to ensure that there was no more than 1 mismatch at greater than

1% within the SARS-CoV-2 database (10,151 sequences). The following table provides the output of the analysis.

| QuantuMDx name | Mismatch rates of single base |
|---|---|
| Orf1_Span20_FPrimer | (27) = 0.27% |
| Orf1_Span20_Probe1 | (23) = 0.23% |
| Orf1_Span20_RPrimer | (0) = 0.0% |
| sGene_Span3_Fprimer | (11) = 0.11% |
| sGene_Span3_Probe | (1) = 0.01% |
| sGene_Span3_Rprimer | (0) = 0.0% |
| nGene_Span1_Fprimer | (29) = 0.29% |
| nGene_Span1_Probe | (6) = 0.06% |
| nGene_Span1_Rprimer | (0) = 0.0% |

Based upon the *in silico* analysis the selected primers and probes will detect all known sequences (as of the date of analysis) of SARS-CoV-2.

#### 24. EXCLUSIVITY (CROSS-REACTIVITY)

The Q-POC™ SARS-CoV-2 Assay primer and probe sequences were used for *In silico* exclusivity testing against 70 micro-organisms (Table 1 and Table 2) identified as potential interfering organisms. The table below provides the lists of organisms utilized for the exclusivity or cross-reaction analysis.

| Organism | Type | Count | Sequence Type |
|---|---|---|---|
| Human coronavirus 229E | Virus | 100 (Limited) | Any |
| Human coronavirus OC43 | Virus | 100 (Limited) | Any |
| Human coronavirus HKU1 | Virus | 100 (Limited) | Any |
| Human coronavirus NL63 | Virus | 100 (Limited) | Any |
| SARS-coronavirus | Virus | 100 (Limited) | Any |
| MERS-coronavirus | Virus | 100 (Limited) | Any |
| Adenovirus (e.g., C1 Ad. 71) | Virus | 100 (Limited) | Any |
| Human Metapneumovirus (hMPV) | Virus | 100 (Limited) | Any |
| Parainfluenza virus 1-4 | Virus | 29 | Any |
| Influenza A & B | Virus | 100 (Limited) | Complete Genomes |
| Enterovirus (e.g., EV68) | Virus | 17 | Complete Genomes |
| Respiratory syncytial virus | Virus | 1 | Complete Genome |
| Rhinovirus | Virus | 4 | Complete Genomes |
| Chlamydia pneumoniae | Virus | 5 | Complete Genomes |
| Haemophilus influenzae | Prokaryote | 1 | Reference Genome |
| Legionella pneumophila | Prokaryote | 1 | Reference Genome |
| Mycobacterium tuberculosis | Prokaryote | 1 | Reference Genome |
| Streptococcus pneumoniae | Prokaryote | 1 | Reference Genome |
| Streptococcus pyogenes | Prokaryote | 1 | Reference Genome |
| Bordetella pertussis | Prokaryote | 1 | Reference Genome |
| Mycoplasma pneumoniae | Prokaryote | 1 | Reference Genome |
| Pseudomonas aeruginosa | Prokaryote | 1 | Reference Genome |
| Staphylococcus epidermis | Prokaryote | 1 | Reference Genome |
| Staphylococcus salivarius | Prokaryote | 1 | Reference Genome |
| Pneumocystis jirovecii (PJP) | Eukaryote | 1 | Reference Genome |
| Candida albicans | Eukaryote | 1 | Reference Genome |
| Human | Eukaryote | 1 | Reference Genome |
| Bat Betacoronavirus | Virus | 100 (Limited) | Any |

| | | | |
|---|---|---|---|
| Bocavirus | Virus | 74 | Any |
| Coronaviridae | Virus | 100 (Limited) | Any |
| Coronavirinae | Virus | 4 | Any |
| Cytomegalovirus | Virus | 1 | Any |
| Enterovirus A | Virus | 1 | Reference Genome |
| Enterovirus B | Virus | 1 | Reference Genome |
| Enterovirus C | Virus | 1 | Reference Genome |
| Enterovirus D | Virus | 1 | Reference Genome |
| Enterovirus E | Virus | 1 | Reference Genome |
| Enterovirus F | Virus | 1 | Reference Genome |
| Enterovirus G | Virus | 1 | Reference Genome |
| Enterovirus H | Virus | 1 | Reference Genome |
| Enterovirus I | Virus | 1 | Reference Genome |
| Enterovirus J | Virus | 1 | Reference Genome |
| Enterovirus K | Virus | 1 | Reference Genome |
| Enterovirus L | Virus | 1 | Reference Genome |
| Human gammaherpesvirus 4 (Epstein-Barr Virus) | Virus | 1 | Reference Genome |
| Human herpesvirus 1 | Virus | 1 | Reference Genome |
| Human herpesvirus 2 | Virus | 1 | Reference Genome |
| Lymphocytic choriomeningitis virus - Segment S | Virus | 1 | Reference Genome |
| Lymphocytic choriomeningitis virus – Segment L | Virus | 1 | Reference Genome |
| Measles | Virus | 1 | Reference Genome |
| Acinetobacter baumannii | Prokaryote | 1 | Reference Genome |
| Bacillus anthracis | Prokaryote | 1 | Reference Genome |
| Chlamydia psittaci | Prokaryote | 1 | Reference Genome |
| Corynebacterium diphtheriae | Prokaryote | 1 | Reference Genome |
| Corynebacterium sp. | Prokaryote | 1 | Reference Genome |
| Coxiella burnetii | Prokaryote | 1 | Reference Genome |
| Enterobacter cloacae | Prokaryote | 1 | Reference Genome |
| Escherichia coli | Prokaryote | 1 | Reference Genome |
| Haemophilus parainfluenzae | Prokaryote | 1 | Reference Genome |
| Klebsiella pneumoniae | Prokaryote | 1 | Reference Genome |
| Mycoplasma hominis | Prokaryote | 1 | Reference Genome |
| Mycoplasma hyorhinis | Prokaryote | 1 | Reference Genome |
| Mycoplasma synoviae | Prokaryote | 1 | Reference Genome |
| Neisseria sicca | Prokaryote | 1 | Reference Genome |
| Proteus vulgaris | Prokaryote | 1 | Reference Genome |
| Staphylococcus aureus | Prokaryote | 1 | Reference Genome |
| Streptococcus GrpA: gallolyticus (firmicutes) | Prokaryote | 1 | Reference Genome |
| Aspergillus fumigatus | Eukaryote | 1 | Reference Genome |
| Candida glabrata | Eukaryote | 1 | Reference Genome |
| Cryptococcus neoformans | Eukaryote | 1 | Reference Genome |

Upon analysis completion, no primers or probes met the criteria of greater than 75% homology, indicating by *in silico* analysis there is minimal chance of off target signal generation.

*In vitro* analysis for exclusivity or cross-reaction was also undertaken. The table below provides the list of organisms utilised for the analysis with no cross-reactivity observed under the tested conditions. All micro-organisms were used at a concentration of $1x10^5$ cfu per reaction.

| Organism | Type | DNA/RNA | Positive/Repeats |
|---|---|---|---|
| Adenovirus (Type 7A) | Virus | RNA | 0/5 |
| Human Metapneumovirus | Virus | RNA | 0/5 |
| Parainfluenza virus 1 | Virus | RNA | 0/5 |
| Influenza A | Virus | RNA | 0/5 |
| Influenza B | Virus | RNA | 0/5 |
| Enterovirus A-L (AMPLIRUN® ENTEROVIRUS 68 RNA CONTROL) | Virus | RNA | 0/5 |
| Respiratory syncytial virus A | Virus | RNA | 0/5 |
| Respiratory syncytial virus B | Virus | RNA | 0/5 |
| Rhinovirus | Virus | RNA | 0/5 |
| Chlamydia pneumoniae | Prokaryote | DNA | 0/5 |
| Haemophilus influenzae | Prokaryote | DNA | 0/5 |
| Legionella pneumophila | Prokaryote | DNA | 0/5 |
| Mycobacterium tuberculosis | Prokaryote | DNA | 0/5 |
| Streptococcus pneumoniae | Prokaryote | DNA | 0/5 |
| Streptococcus pyogenes | Prokaryote | DNA | 0/5 |
| Bordetella pertussis | Prokaryote | DNA | 0/5 |
| Mycoplasma pneumoniae | Prokaryote | DNA | 0/5 |
| Pneumocystis jirovecii | Eukaryote | DNA | 0/5 |
| Pooled human nasal wash* (*see Human Coronavirus) | Not Applicable | NA | 0/5 |
| Candida albicans | Eukaryote | DNA | 0/5 |
| Pseudomonas aeruginosa | Prokaryote | DNA | 0/5 |
| Staphylococcus epidermis | Prokaryote | DNA | 0/5 |
| Streptococcus salivarius | Prokaryote | DNA | 0/5 |
| Human coronavirus 229E | Virus | RNA | 0/5 |
| Human coronavirus OC43 | Virus | RNA | 0/5 |
| Human coronavirus*<br>In place of evaluating pooled human nasal wash, testing of a pool of 10 individual negative clinical swab specimens was performed to represent diverse microbial flora in the human respiratory tract HKU1 | Virus | RNA | 0/5 |
| Human coronavirus NL63 | Virus | RNA | 0/5 |

#### 25. INTERFERING SUBSTANCES

Potential interfering substances that could be present in an upper respiratory specimen were evaluated. The substances listed include both endogenous as well as exogenous substances. None of the tested substances under the conditions tested showed an ability to interfere with the detection of SARS-CoV-2.

| Interfering Substance | Concentration tested | Positive/Repeats |
|---|---|---|
| Mucin – bovine submaxillary gland, type I-S | 5 mg/mL | 5/5 |
| Blood (Human) | 0.25% (v/v) | 5/5 |
| Phenylephine hydrochloride, Max stregth Cold and Flu Relief, Boots | 0.2 mg/mL | 5/5 |
| Oxymetazoline hydrochloride, Blocked nose relief, Boots | 30% (v/v) | 5/5 |
| NasalGuard Cold and Flu block | 40 mg/mL | 5/5 |
| Galphimia glauca, cardiospermum and Luffa operculata, Rinital (25mg/tablet) | 12.5 mg/mL | 5/5 |
| Benzocaine, Orajel Dental gel | 2.5 mg/mL | 5/5 |
| Menthol, Lockets | 0.084% (w/v) | 5/5 |
| Zanamivir, antiviral drug | 5 mg/mL | 5/5 |
| Beclomethanzone diproprionate | 5 mg/mL | 5/5 |
| Mupirocin | 1.2 mg/mL | 5/5 |
| Tobramycin | 0.005% (v/v) | 5/5 |

The performance of this assay has not been established in patients receiving intranasal administered influenza vaccine. The performance of this assay has not been established in immunocompromised individuals.

#### 26. REPRODUCIBILITY

The Q-POC™ SARS-CoV-2 Assay was evaluated at 3x LoD for day-to-day, tech-to-tech, instrument-to-instrument, site-to-site and lot-to-lot for reproducibility and repeatability under the conditions tested. Across all evaluations, reproducibility was >99%.

#### 27. CLINICAL PERFORMANCE

A clinical evaluation was performed using SARS-CoV-2 suspected specimens. Results for the Q-POC™ SARS-CoV-2 Assay are shown below against the QuantuMDx SARS-CoV-2 RT-PCR Detection Assay. A Ct cut-off value of 35 was set on the QuantuMDx SARS-CoV-2 RT-PCR Detection Assay. The clinical performance study was carried out at two sites, QuantuMDx and St George's University Hospitals (SGUL).

| | | QuantuMDx SARS-CoV-2 RT-PCR Detection Assay | |
|---|---|---|---|
| | | Positive | Negative |
| Q-POC™ SARS-CoV-2 | Positive | 31 | 2 |
| | Negative | 1 | 113 |

**PPA (clinical sensitivity): 96.9% (95% CI: 83.8% - 99.9%)**
**NPA (clinical specificity): 98.3% (95% CI: 93.9% - 99.8%)**
**98.0% agreement across all samples**

#### 28. DISPOSAL

Used test cassettes should be considered biohazardous.
Dispose of all hazardous or biologically contaminated materials in accordance with local, regional and national laws and in accordance with the practices of your institution.

#### 29. REFERENCES

- Center for Disease Control and Prevention. Biosafety in Microbiological and Biomedical Laboratories, 5th ed. U.S. Department of Health and Human Services, Public Health Service. Centers for Disease Control and Prevention, National Institute of Health HHS Publication No. (CDC) 21-1112, revised December 2009.
- Clinical and Laboratory Standards Institute (CLSI). Protection of laboratory workers from occupationally acquired infections. Approved Guideline – Fourth Edition. CLSI Document M29-A4; Wayne, PA.
- Clinical and Laboratory Standards Institute (CLSI). Collection, Transport, Preparation and Storage of Specimens for Molecular Methods Approved Guideline – First Edition. CLSI Document M13-A; Wayne, PA.
- World Health Organization. Laboratory Biosafety Manual, 3rd ed. Geneva Switzerland; World Health Organization; 2004
- Center for Disease Control and Prevention. Interim Guidelines for Collecting, handling and Testing Clinical Specimens from Persons Under Investigation (PUIs) for Coronavirus Disease 2019 (CoVID-19). Available at – https://www.cdc.gov/coronavirus/2019-nCoV/lab/guidelines-clinical -specimens.html
- Clinical and Laboratory Standards Institute (CLSI). Statistical Quality Control for Quantitative Measurements: Principles and Definitions Approved Guideline – Second Edition. CLSI Document C24-A2; Wayne, PA.

#### 30. SYMBOLS

| Symbol | Interpretation | Symbol | Interpretation |
|---|---|---|---|
| | Consult instructions | | Important information. Please read carefully |
| | Used for both warnings and cautions.<br>A warning indicates the risk of personal injury or loss of life if the operating procedures and practices are not correctly followed.<br>A caution indicates the possibility of loss of data or damage to, or destruction of equipment if operating procedures and practices are not strictly observed | | Biohazard: Follow proper infection control guidelines for handling all specimens and samples. Properly dispose of all contaminated waste according to local requirements |
| | Indicates the product's temperature limits | CONTROL + | Indicates Positive Control material |
| | Indicates Use by Date | CONTROL – | Indicates Negative Control Material |
| LOT | Indicates the product batch code | | Fragile |
| | Indicates the name and location of the product manufacturer | | Do not use if package is damaged |
| REF | Indicates the product's catalogue number | Σ n | Indicates the kit contents sufficient for <n> tests |
| | Indicates single use only. Do not reuse | CE IVD | Product is CE marked in compliance with Directive 98/79/EC as an in vitro diagnostic medical device |
| | Indicates product's humidity limits | Rx Only | Product is for prescription use only |

### 31. CONTACT INFORMATION

For customer support please contact QuantuMDx directly. Information can be found at:

QuantuMDx Group Ltd.
Lugano Building
57 Melbourne Street
Newcastle upon Tyne
United Kingdom
NE1 2JQ

Website: quantumdx.com
Telephone hours: 9 – 5 (GMT)

Customer Support Email:

Order Support Email:
